# Plasma phosphorylated tau 217 detects amyloid-β in Neuronal Synuclein Disease

**DOI:** 10.1101/2025.09.07.25335217

**Authors:** Alena M. Smith, Sara A. Lorkiewicz, Burak Arslan, Laia Montoliu-Gaya, Nicholas J. Ashton, Edward N. Wilson, Daniel Alcolea, Íñigo Rodríguez-Baz, Juan Fortea, Christina B. Young, Joseph R. Winer, Marian Shahid-Besanti, Hillary Vossler, Melanie J. Plastini, Tianyu Pan, Elena Vera-Campuzano, Isabel Sala, Justin H. Mendiola, Veronica Ramirez, Geoffrey A. Kerchner, Katrin I. Andreasson, Victor W. Henderson, Thomas J. Montine, Lu Tian, Elizabeth C. Mormino, Henrik Zetterberg, Kathleen L. Poston, Carla Abdelnour

## Abstract

**Background:** Multiple proteinopathies commonly coexist in neurodegenerative diseases, therefore it is critical to understand plasma biomarker performance to detect proteinopathies in these complex diseases. While plasma biomarkers can accurately detect amyloid-β in individuals with Alzheimer’s disease, their performance accuracy is unknown in individuals with Neuronal Synuclein Disease (NSD).

**Objective:** To determine the accuracy of plasma pTau217, pTau181, Aβ42/40, GFAP, and NfL to detect amyloid-β in NSD. Additionally, to establish and validate cut-points for the most promising amyloid-β plasma biomarker in NSD.

**Methods:** This cross-sectional cohort study analyzed data from two observational cohorts from Stanford University and Sant Pau Hospital. The discovery cohort participants were biologically-defined by NSD and Aβ status among clinical Lewy body disease (LBD), Alzheimer’s disease (AD), or cognitively unimpaired (CU) individuals. The two validation cohorts consisted of clinically-defined LBD participants. Data were analyzed from 2/2024-3/2025. Plasma pTau217, pTau181, Aβ42/40, GFAP, and NfL were analyzed as potential biomarkers for amyloid-β in NSD, using cerebrospinal fluid Aβ42/40 and Aβ positron emission tomography as reference gold-standards. Diagnostic accuracy was determined using Receiver Operating Characteristic (ROC) analysis.

**Results:** We included 253 participants (mean[SD] age=71[9.9] years), 180 from the discovery cohort and 73 from the clinical validation cohorts. In the discovery cohort, plasma pTau217, pTau181, Aβ42/40, and GFAP levels significantly differed between Aβ+ and Aβ-participants, regardless of NSD status. Plasma NfL levels were significantly higher in the NSD+/Aβ+ group compared to all other groups. Plasma pTau217 showed the largest median fold-change between Aβ+ and Aβ-participants and demonstrated the highest diagnostic performance in detecting amyloid-β in NSD (Area Under the Curve=0.92, 95% CI=0.81-0.98). Applying one-or two-reference cut points for plasma pTau217 in the validation cohorts could reduce the need for additional amyloid-β testing in 41-56% of clinically-defined LBD participants.

**Conclusions:** Plasma pTau217 accurately detects amyloid-β in NSD individuals, with reproducible cut points in clinical LBD cohorts. Our findings demonstrate plasma pTau217 is a cost-effective and minimally-invasive tool for determining amyloid-β in mixed-etiology neurodegenerative diseases of aging.

## Introduction

In neurodegenerative diseases of aging, coexistence of amyloid-β, tau, and α-synuclein (αSyn) is common and contributes to clinical heterogeneity, faster disease progression, and variable treatment response. Advances in biomarker technology now enable for accurate, scalable, and cost-effective detection of some of these proteinopathies. As these biomarkers move into clinical practice, it is essential to understand their performance in the context of mixed neurodegenerative diseases.

Lewy body disease (LBD), which includes Parkinson’s disease (PD) and dementia with Lewy bodies (DLB), is characterized by the abnormal deposition of αSyn. This diagnostic pathologic feature can be detected in cerebrospinal fluid (CSF) using the seed amplification assay (SAA), supporting the biologically based definition of Neuronal Synuclein Disease (NSD)(1).

Concomitant amyloid-β is frequently found in NSD and in clinically diagnosed PD and DLB(2–6), and is associated with worse clinical outcomes(2), earlier dementia onset(7), faster cognitive decline(8,9) and shorter survival(10). These findings underscore the importance of identifying NSD individuals with concomitant amyloid-β, and the potential value of testing anti-amyloid therapies in this population.

Blood-based biomarkers have emerged as promising tools for detecting amyloid-β with accuracy comparable to CSF and positron emission tomography (PET) imaging in Alzheimer’s disease (AD)(11,12). Among them, plasma phosphorylated tau at threonine 217 (pTau217) has consistently outperformed other plasma biomarkers such as pTau181 and the Aβ42/40 ratio for detecting amyloid-β in AD(11,13,14). Combinations of these markers, either with each other or with markers of neuroinflammation (i.e. glial fibrillary acidic protein, GFAP) and neurodegeneration (i.e. neurofilament light chain, NfL), have shown lower performance than pTau217 alone(11). Based on this evidence, pTau217 is now recommended for clinical and research use(15).

Multiple studies have evaluated plasma pTau181(9,14,16–26), pTau217(9,20,27,28), Aβ42/40 ratio(16,18,19,24), GFAP(16,18,19,24,25,29,30), and NfL(16,18,19,21,22,24–27) in clinically diagnosed LBD participants. However, their diagnostic accuracy for detecting amyloid-β have not been evaluated in biologically defined NSD. This distinction is important as plasma biomarkers for amyloid-β are being considered as a routine component of NSD staging(1), particularly since prior studies show AD biomarker cut points can differ between “mixed” NSD plus AD and “pure” AD cases(31–33). Evaluating plasma biomarkers in NSD provides a closer approximation to underlying pathology than clinical diagnosis, which often does not predict Lewy bodies at autopsy(34–36).

As plasma biomarkers enter clinical use, their interpretation must align with the intended use population, as emphasized by the FDA-NIH Biomarker Working(37). In NSD, accurate interpretation of plasma biomarkers is crucial for their appropriate use as screening tools for patient selection and stratification in clinical trials, particularly as drug development begins to address mixed pathologies.

In this study, we assessed whether plasma pTau217, pTau181, Aβ42/40, GFAP, and NfL accurately detect amyloid-β in individuals with biologically defined NSD. We calculated plasma biomarker cut points using one-and two-reference standard approaches, then validated these cut points in two independent clinical LBD cohorts to assess their generalizability. By establishing and validating plasma biomarker cut points for detecting amyloid-β in NSD, this study addresses a critical gap in biomarker interpretation that currently limits their use in mixed pathology settings.

## Material and Methods

### 1. Participants

This cross-sectional study included participants from two cohorts: Stanford University (SU) and the Sant Pau Initiative on Neurodegeneration (SPIN)(38), recruited between 2011 and 2023 (see Supplementary Methods). The Institutional Review Boards of each academic institution approved the study protocols. All participants or their legally authorized representatives provided written informed consent in accordance with the Declaration of Helsinki. This study followed the Strengthening the Reporting of Observational Studies in Epidemiology (STROBE) reporting guidelines.

All participants underwent comprehensive clinical evaluation involving medical history, physical and neurological examination, and neuropsychological assessment. Diagnoses were based on international consensus criteria, similar to our prior studies(9,39) and included: LBD without cognitive impairment (LBD-CU; i.e., PD diagnosis according to UK Brain Biobank criteria(40) without objective impairment on neuropsychological testing), LBD with cognitive impairment (LBD-CI; i.e., diagnosis of PD with mild cognitive impairment(41), prodromal DLB(42), PD dementia(43) or DLB(44)), AD(45), and cognitively unimpaired (CU) individuals (i.e., older adults without parkinsonian symptoms who performed within normal age-and sex-adjusted ranges on neuropsychological testing).

Participants were divided into one discovery and two clinical validation cohorts. The discovery cohort comprised 180 SU participants with CSF αSyn SAA and Aβ42/40 data, who had clinical diagnoses of CU (n=86), LBD-CU (n=30), LBD-CI (n=30) and AD (n=34). Participants were biologically classified into four groups according to CSF αSyn and Aβ status: NSD-/Aβ-, NSD-/Aβ+, NSD+/Aβ-, and NSD+/Aβ+. Within the SU discovery cohort, we evaluated diagnostic accuracy of plasma biomarkers for detecting amyloid-β in NSD+ and NSD-groups.

The two clinical validation cohorts comprised clinically-defined LBD-CU and LBD-CI participants (CSF αSyn SAA data were not available). Amyloid-β status was determined by Aβ PET in the LBD-SU cohort and CSF Aβ42/40 in the LBD-SPIN cohort. The LBD-SU cohort included 29 participants with LBD-CI (n=16) or LBD-CU (n=13), and the LBD-SPIN cohort included only LBD-CI participants (n=44). Within these cohorts, we validated diagnostic accuracies of plasma biomarker cut points defined in the SU discovery cohort (Supplementary Figure 1).

### 2. Biomarkers

All biomarker processing for plasma, CSF, and Aβ PET imaging was performed blinded to clinical information. In SU, CSF samples were obtained within six months of plasma collection (mean [SD]: 2.76 [2.24] weeks) and Aβ PET within one year of plasma collection (mean [SD]: 14.60 [13.85] weeks). In SPIN, CSF samples were obtained at time of plasma collection. Detailed biomarker collection and processing procedures are provided in Supplementary Methods.

### 2.1. Plasma

Plasma pTau181, Aβ42, and Aβ40 concentrations were measured by the Stanford Alzheimer’s Disease Research Center (ADRC) Biomarker Core using the Lumipulse *G*1200 instrument.

Plasma pTau181 was quantified using a modified version of the Lumipulse *G* CSF pTau181 assay (231654; Fujirebio Diagnostics)(46). Plasma Aβ42 and Aβ40 were measured using the Lumipulse *G* plasma Aβ42 and Aβ40 assays(47).

Plasma pTau217, GFAP, and NfL concentrations were measured at the University of Gothenburg Department of Psychiatry and Neurochemistry, as detailed elsewhere(11). Plasma pTau217 was measured with the commercial ALZpath pTau217 assay, and GFAP and NfL with the commercial Neurology 2-plex E kit (103670; Quanterix).

### 2.2. CSF αSyn, Aβ42, and Aβ40

CSF sample collection procedures were similar across SU and SPIN cohorts (Supplementary Methods). CSF αSyn status in the SU discovery cohort was determined at Amprion’s CLIA Laboratory via αSyn seed amplification assay (SYNTap® Biomarker Test)(48). CSF samples were classified as aSyn “detected” (NSD+) or “not detected” (NSD-) based on a preestablished threshold for median maximum fluorescence of the triplicate wells within 150 hours. The complete protocol is published elsewhere(49). SU CSF Aβ42 and Aβ40 were measured using the Lumipulse platform as previously described(50).

### 2.3. Aβ PET

We acquired ^18^F-florbetaben imaging on a PET/MRI scanner (Signa 3T, GE Healthcare) at the Stanford University Richard M. Lucas Center for Imaging. Stanford ADRC Aβ PET acquisition parameters were previously published(51) and are briefly described in the Supplementary Methods.

CSF Aβ status was determined using Aβ PET as the references stardard in the SU discovery cohort (Supplementary Methods). Aβ PET positivity was defined using a threshold of 36 centiloids (CL) to maximize specificity (Supplementary Figure 2). This threshold was derived from a subset of SU participants excluded from discovery and validation cohorts using a Gaussian Mixture Model (Supplementary Methods). In an exploratory analysis, we used a 24 CL threshold(11) to maximaze sensitivity (Supplementary Table 1). Both 36 and 24 CL Aβ PET thresholds were used to establish CSF Aβ42/40 cut points calculated with the Youden Index in ROC curve analyses, where abnormal CSF Aβ42/40 was defined as < 0.09 for 36 CL and < 0.11 for 24 CL. With these CSF Aβ42/40 cut points, we classified 180 SU participants as either CSF Aβ+ or CSF Aβ-for analysis.

### 3. Statistical analysis

Plasma pTau217, pTau18, GFAP, and NfL data were log10-transformed due to skewed distributions. We summarized continuous variables using the mean (standard deviation) for normally distributed data, median (minimum, maximum) for non-normally distributed data, and proportions for categorical variables.

To assess group differences in continuous variables, we used either one-way ANOVAs or the Kruskal-Wallis tests, depending on data distribution. We performed post-hoc pairwise comparisons using *t*-tests or Mann-Whitney tests as appropriate. For categorical variables, we used either Pearson’s χ^2^ or Fisher’s Exact tests depending on observed proportions. We corrected for multiple comparisons using the Holm method.

To quantify the differences in plasma biomarker levels between Aβ+ and Aβ-participants in the SU discovery cohort, we calculated median fold-change, standard deviation, and 95% confidence intervals. With this cohort, we conducted univariable and multivariable linear regression analyses to evaluate associations between plasma biomarker levels and age, sex, *APOE* ε4 carriership (i.e., ε4 homozygotes or heterozygotes), CSF Aβ, and NSD status separately and jointly.

To determine diagnostic accuracy of plasma biomarkers for detecting amyloid-β in NSD+ and NSD-, we used Receiver Operating Characteristic (ROC) curves. First, we analyzed each biomarker separately, incorporating age, sex, and *APOE* ε4 carriership in each model. Next, we combined multiple biomarkers into a single model. Model performance was compared with the DeLong test, and the best-performing biomarker was selected based on standard metrics. For this biomarker, we determined one-reference cut point using the Youden Index, as well as two-reference cut points based on 90% and 95% sensitivity and specificity. Then, we calculated the predicted probabilities for amyloid-β posititivy using logistic regression under the two-reference models, and stratified participants into low, intermediate, and high-risk. Finally, we calculated sensitivity, specificity, negative and positive percent agreement, as well as overall agreement.

Statistical analyses were performed using R version 4.3.0 (R Foundation for Statistical Computing). Two-sided *p*-values ≤ 0.05 were statistically significant and 95% confidence intervals were reported when applicable.

## Results

### 1. Participants characteristics

The SU discovery cohort (n=180) had a median age of 69 years (min, max: 50-87), with 83 females (46.1%) and 97 males (53.9%). Participants were classified into four biologically defined groups: NSD-/Aβ-(n=69), NSD-/Aβ+ (n=44), NSD+/Aβ-(n=39), and NSD+/Aβ+ (n=28). The NSD-/Aβ-group was younger than the NSD-/Aβ+ group. Otherwise, the groups were similar in age, sex, and years of education (Table 1).

**Table 1.**
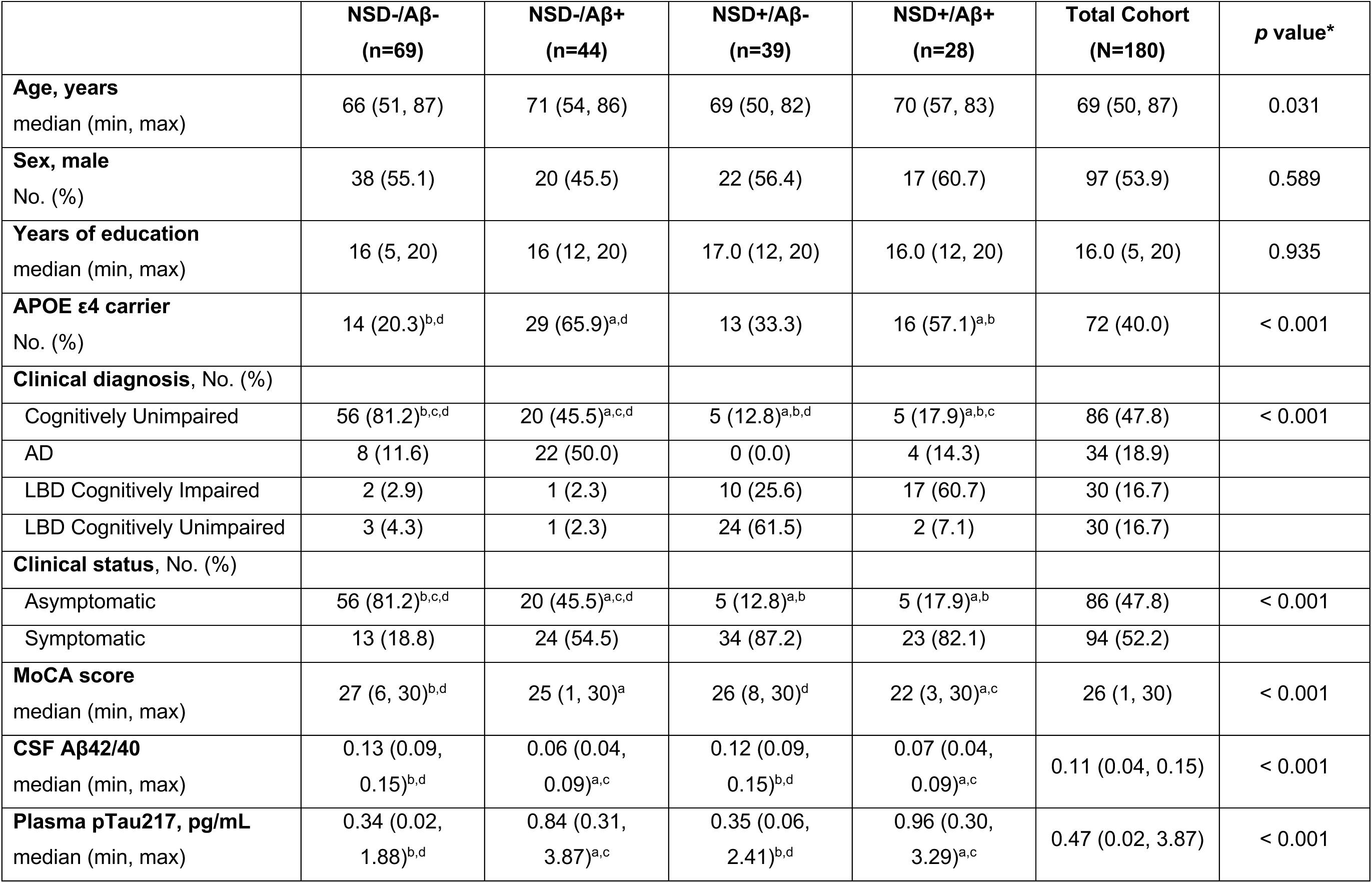
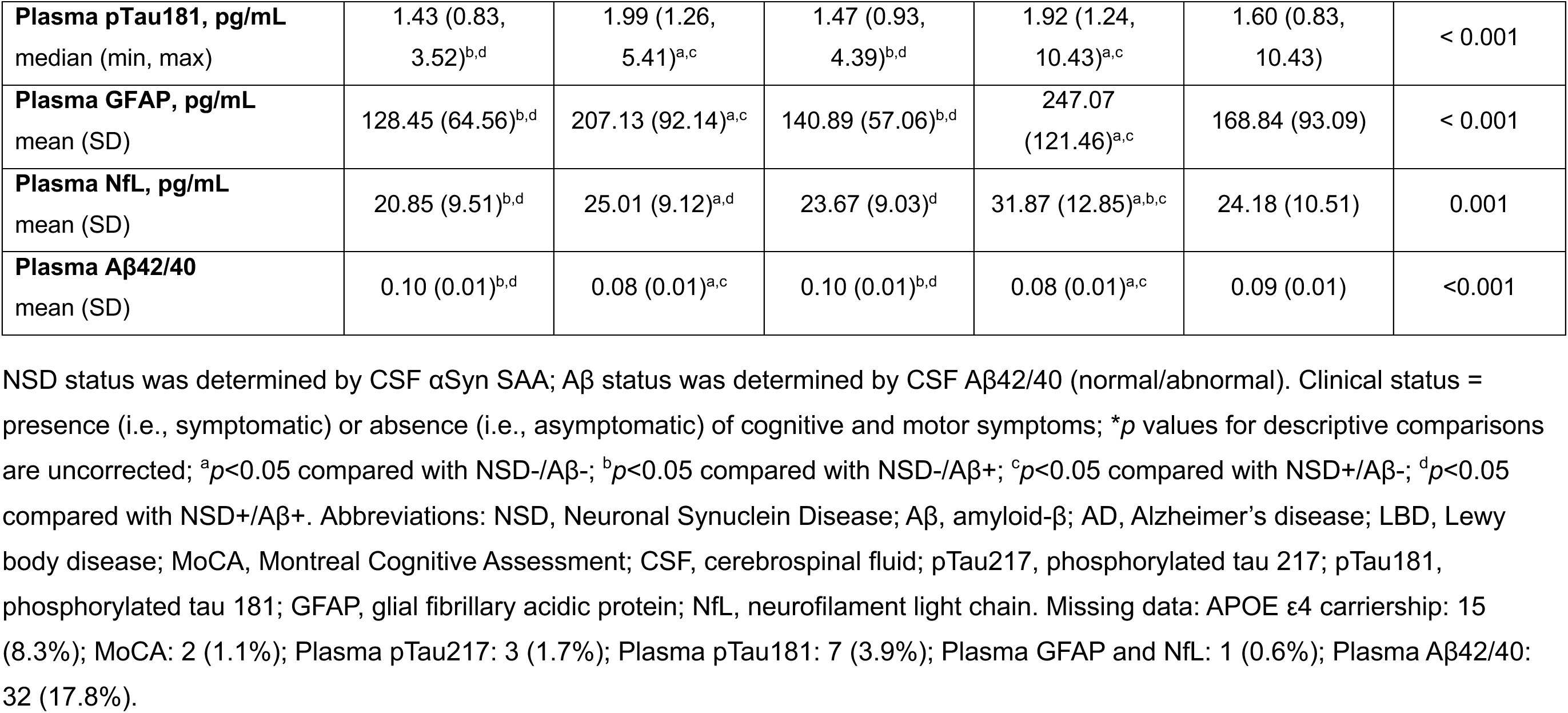
Characteristics of the Stanford University discovery cohort.

The LBD-SU cohort had a median age of 71 years (min, max: 57-81), with 6 females (20.7%), and 23 males (79.3%). The LBD-SPIN cohort had a median age of 78 years (min, max: 65-84), with 20 females (45.5%) and 24 males (54.5%) (Supplementary Table 2).

### 2. Plasma biomarker levels stratified by CSF NSD and Aβ status

In the SU discovery cohort, plasma levels of pTau217, pTau181, and GFAP were significantly higher, while Aβ42/40 was significantly lower in the Aβ+ compared to the Aβ-groups, regardless of NSD status (Figure 1, panels A-D; Table 1). Plasma NfL levels were significantly higher in the NSD+/Aβ+ group compared to all other groups, and in the NSD-/Aβ+ compared to the NSD-/Aβ-group (Figure 1, panel E; Table 1).

**Figure 1.**
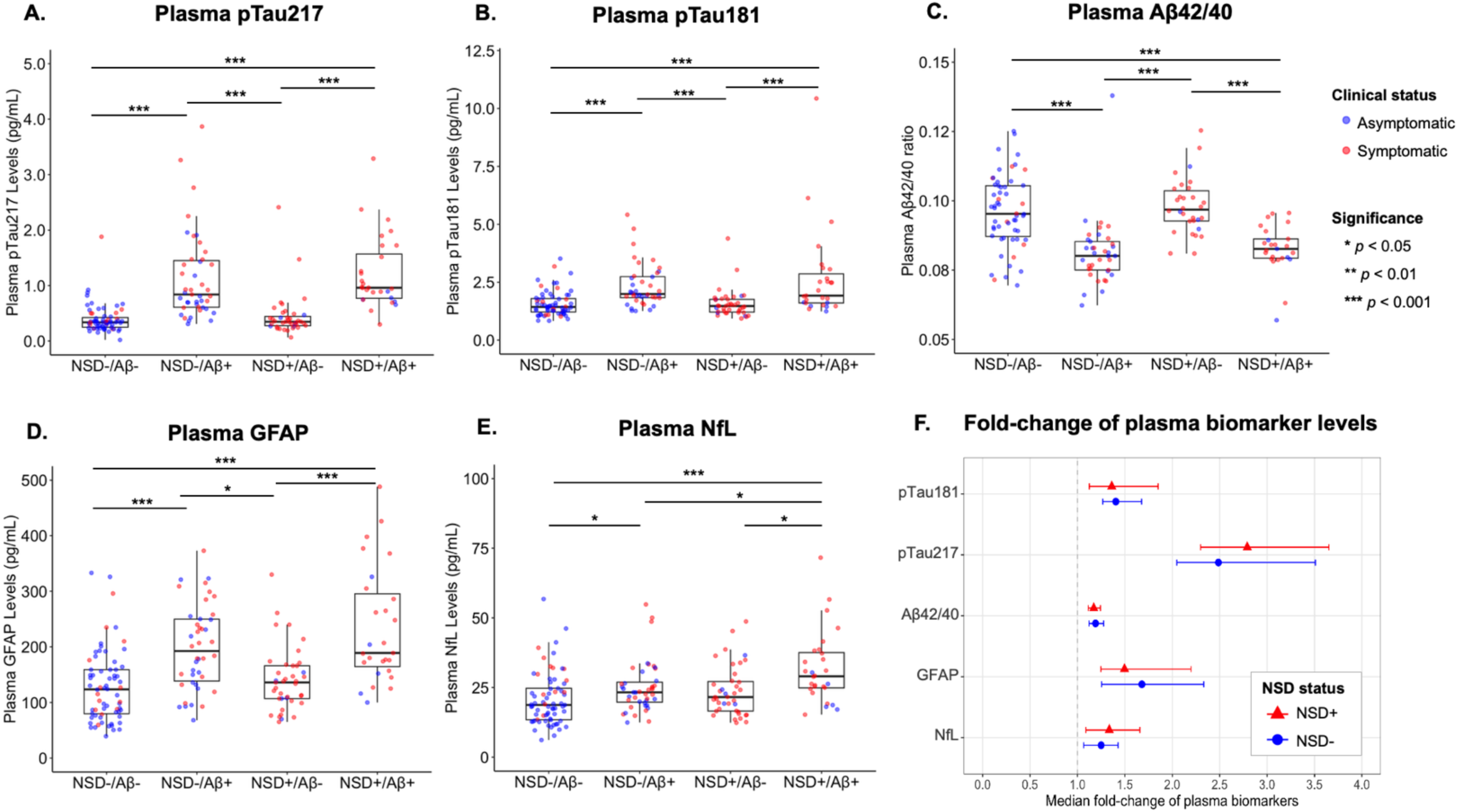
Plasma biomarker levels (pg/mL) across NSD/Aβ groups. Panel 1A-1E: Bars with asterisks represent significant differences between log-transformed plasma biomarker levels. Plasma pTau217, pTau181, Aβ42/40, and GFAP levels were abnormal in Aβ+ groups, regardless of NSD status. Plasma NfL levels were higher in the NSD+/Aβ+ group compared to the other groups; and in the NSD-/Aβ+ group compared to the NSD/Aβ-group. Plasma pTau217 showed the largest median fold-change regardless of NSD status, with the highest median concentration observed in the NSD+ group (2.79 [SD: 0.33], 95% CI = 2.30-3.61; see Supplementary Table 4).

Visual inspection identified one outlier in the NSD+/Aβ+ group with a pTau181 level of 10.43 pg/mL (7.9 SDs above the mean). Removing this outlier in a sensitivity analysis did not change the results. Another sensitivity analysis excluding symptomatic participants (presenting with cognitive and/or motor symptoms) from the NSD-/Aβ-group, and asymptomatic participants from the NSD-/Aβ+, NSD+/Aβ+, and NSD+/Aβ-groups showed consistent findings except for plasma NfL (Supplementary Figure 3), suggesting that symptom severity might be associated with NfL levels.

Figure 1-panel F shows the median fold-change of plasma biomarkers in the SU discovery cohort. Plasma pTau217 showed the largest median fold-change between Aβ+ and Aβ-participants, with the highest value in the NSD+ group (2.79 [SD: 0.33], 95% CI = 2.3-3.61; Supplementary Table 3).

### 3. Association of plasma biomarkers with age, sex and APOE ε4 carriership

Linear regression models examined the effects of age, sex, *APOE* ε4 carriership, CSF Aβ and NSD status on plasma biomarker levels (Supplementary Table 4). In both univariable and multivariable linear regression analyses, higher levels of pTau217 and pTau181 were associated with CSF Aβ+. These results remained consistent after excluding the pTau181 outlier. Lower Aβ42/40 was associated with both *APOE* ε4 carriership and CSF Aβ+.

Additionally, higher GFAP levels were associated with older age, CSF Aβ+, and NSD+, while male sex was associated with lower GFAP levels. Finally, higher NfL levels were associated with older age, CSF Aβ+, and NSD+.

### 4. Diagnostic accuracy of plasma biomarkers for detecting amyloid-β

In NSD+ participants, plasma pTau217 and Aβ42/40 showed the highest accuracy for detecting amyloid-β (Figure 2, panel A). Their diagnostic performances outperformed NfL, but were comparable to pTau181 and GFAP. Including age, sex, and *APOE* ε4 carriership did not improve model accuracies (Supplementary Table 5). Sensitivity analyses confirmed that excluding the pTau181 outlier did not alter its diagnostic performance. However, pTau217 became significantly more accurate than pTau181 after this exclusion.

**Figure 2.**
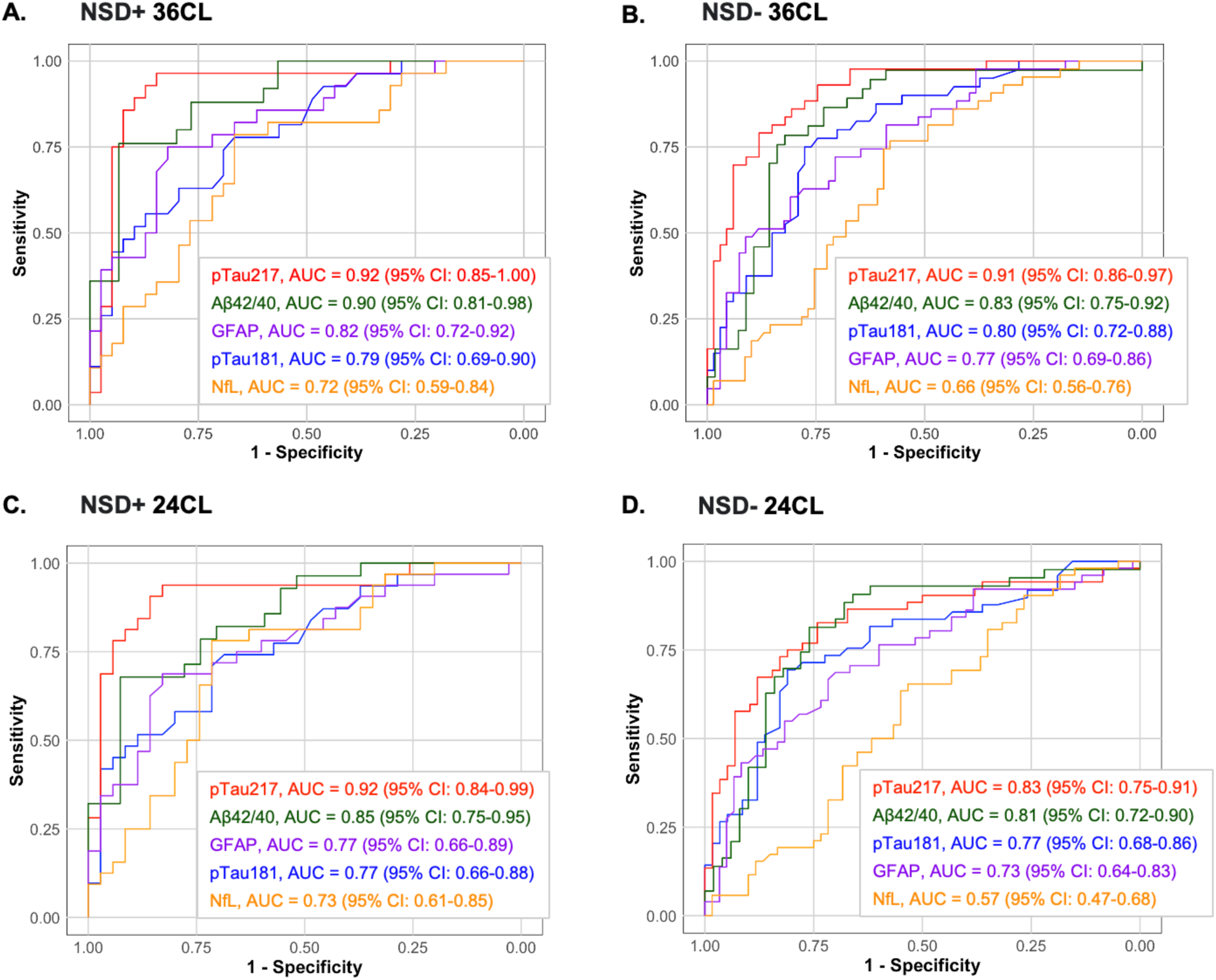
ROC models for detecting amyloid-β using individual plasma biomarkers in NSD+ and NSD-participants. Figure 2 compares diagnostic accuracies of individual plasma biomarker receiver operating characteristic (ROC) models for detecting amyloid-β. Panels 2A and 2C show ROC area under the curve (AUC) results for NSD+ participants whereas panels 2B and 2D show ROC AUC results for NSD-participants. Panels 2A and 2B show models using a CSF Aβ42/40 cut point calculated based on Aβ PET > 36 centiloids (CL). Panels 2C and 2D show models using a CSF Aβ42/40 cut point calculated based on Aβ PET > 24 CL. Plasma pTau217 outperforms other biomarkers for the detection of amyloid-β in NSD+ participants. Individual biomarker models performed similarly when detecting amyloid-β in NSD-participants, with no biomarker significantly outperforming the other.

In NSD-participants, plasma pTau217 and Aβ42/40 again demonstrated the highest accuracies for detecting amyloid-β, with no significant difference between them (Figure 2, panel B). Plasma pTau217 outperformed pTau181, GFAP, and NfL. Including age, sex, and *APOE* ε4 carriership significantly improved NfL’s accuracy, but was still less accurate than pTau217, GFAP, or Aβ42/40 alone.

In both NSD+ and NSD-groups, models incorporating all biomarkers achieved the highest accuracy, but their performance was not significantly better than pTau217 or Aβ42/40 alone (Supplementary Tables 5 and 6).

In an exploratory analysis using an Aβ PET positivity threshold of 24 CL (Figure 2, panels C and D), key differences emerged. In NSD+ participants, pTau217 remained the most accurate biomarker, significantly outperforming pTau181, GFAP, and NfL. In NSD-, all biomarkers performed similarly, except for NfL, which showed lower accuracy. Adding age, sex, and *APOE* ε4 carriership significantly improved NfL’s performance, while other biomarkers showed no significant improvement. In combined biomarker models, the only difference from the 36 CL Aβ PET positivity threshold analyses was in NSD+ participants, where pTau217 performed comparably to all optimal biomarker combinations (Supplementary Tables 7 and 8).

### 5. Validation of plasma pTau217 in two independent clinical LBD cohorts

Plasma pTau217 and Aβ42/40 demonstrated similar accuracy for detecting amyloid-β (Supplementary Tables 5 and 6). However, pTau217 showed the best overall performance in both NSD+ and NSD-groups, with the largest median fold-change between Aβ+ and Aβ-participants. Consequently, we selected pTau217 for further validation of reference cut points for identifying amyloid-β in NSD.

In NSD+ participants from the SU discovery cohort, we established one-reference cut point for plasma pTau217 using the Youden Index (>0.54 pg/mL). We then derived two-reference cut points: one at 90% sensitivity (<0.66 pg/mL) and specificity (>0.69 pg/mL), and another at 95% sensitivity (<0.47 pg/mL) and specificity (>1.04 pg/mL). These cut points differed in NSD-participants (Supplementary Table 9).

Using logistic regression, we calculated predicted probabilities of amyloid-β positivity under the two-reference model, stratifying participants into low, intermediate and high-risk categories (Figure 3, panel A). In the SU discovery cohort, NSD+ participants with a predicted probability <6% had a 95% chance, and those <36% had a 90% chance, of being Aβ-(low probability). Conversely, probabilities >94% had a 95% chance, and >53% had a 90% chance, of being Aβ+ (high probability). Participants with probabilities between 36% and 53% were classified as intermediate risk.

**Figure 3.**
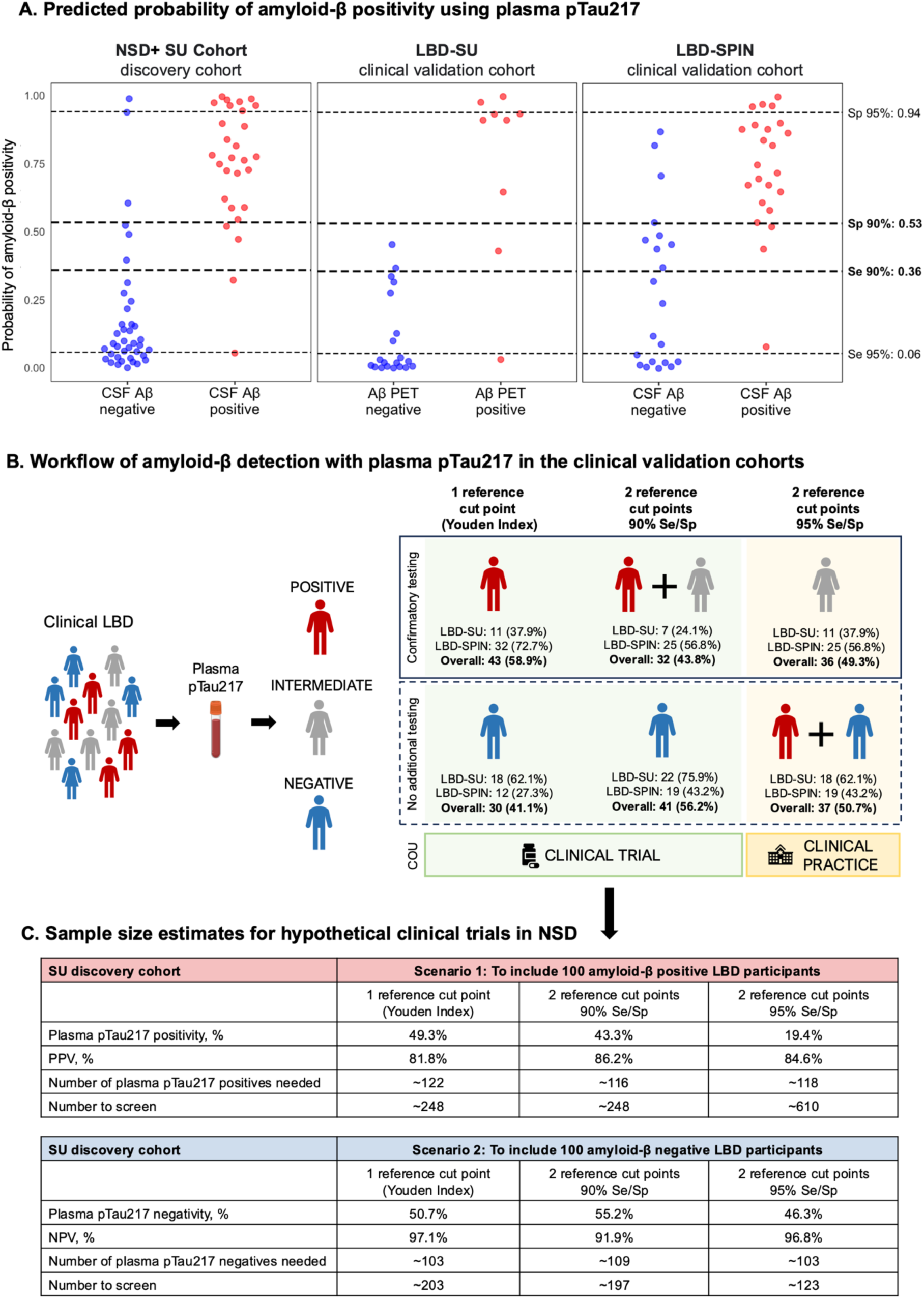
Validation of plasma pTau217 in clinical LBD cohorts. Panel 3A. shows the predicted probability of amyloid-β positivity based on a logistic regression model including z-transformed plasma pTau217 levels across cohorts. Each dot represents an individual participant, with blue dots indicating amyloid-β negative and red dots indicating amyloid-β positive. Predicted probabilities are shown for the NSD+ SU discovery cohort (left). Dashed lines indicate the probability thresholds corresponding to 90% (thick line) and 95% (thin line) sensitivity or specificity, demonstrating low-and high-risk cut points on the probability distribution, applying a similar approach as Brum et al. (doi: 10.1038/s43587-023-00471-5). **Panel 3B** shows a workflow of amyloid-β detection with plasma pTau217 in the clinical validation cohorts. On the left, the flowchart presents the categorization of patients based on their plasma pTau217 levels: positive, intermediate, or negative results. The right panel presents the workflow using different reference cut points. The top section presents the number and percentage of patients that will require confirmatory testing (with PET or CSF), while the bottom section presents the number and percentage of patients that will not require additional testing. The color-coded boxes indicate a potential context of use (COU) for the different reference cut points: clinical trial (green) and clinical practice (yellow), assuming in clinical trials would bias towards more confirmatory testing for those suspected of being amyloid-β positive. **Panel 3C** shows sample size estimates for hypothetical clinical trials, which are calculated using the NSD+ SU discovery cohort. In Scenario 1 the goal was to find 100 NSD who were amyloid-β positive, versus Scenario 2 where the goal was to find 100 NSD who were amyloid-β negative. The number of plasma pTau217 positives (or negatives) needed was calculated as 100 divided by the PPV(or NPV). The number to screen was calculated as plasma pTau217 positives (or negatives) needed divided by the percentage of plasma pTau217 positivity (or negativity).

We applied NSD+ reference cut points to the LBD-SU and LBD-SPIN clinical cohorts (Table 2). The negative percentage agreement (NPA) was similar between the one-and two-reference cut point models at 90% sensitivity/specificity, ranging from 89.5% to 94.4%.

**Table 2.**
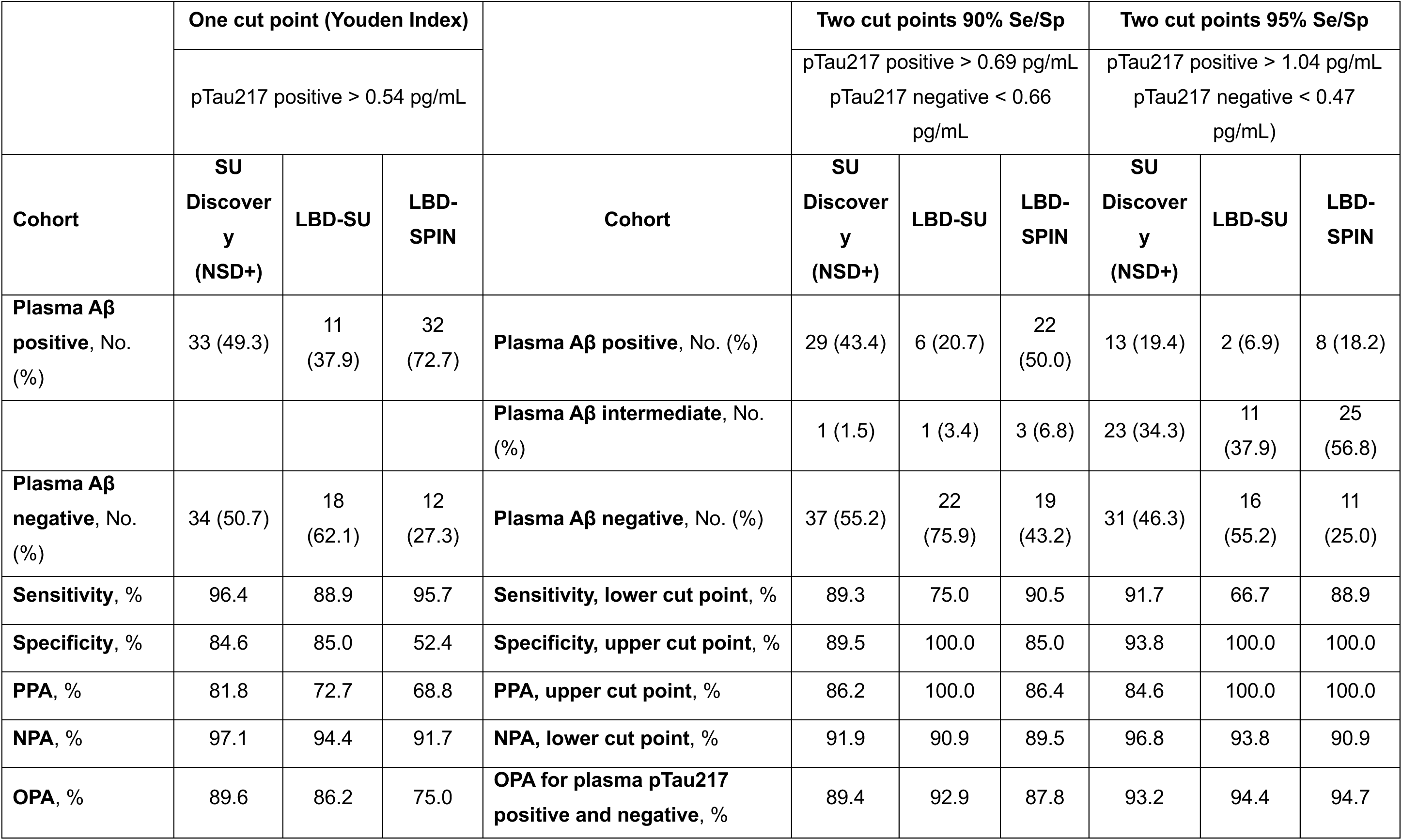
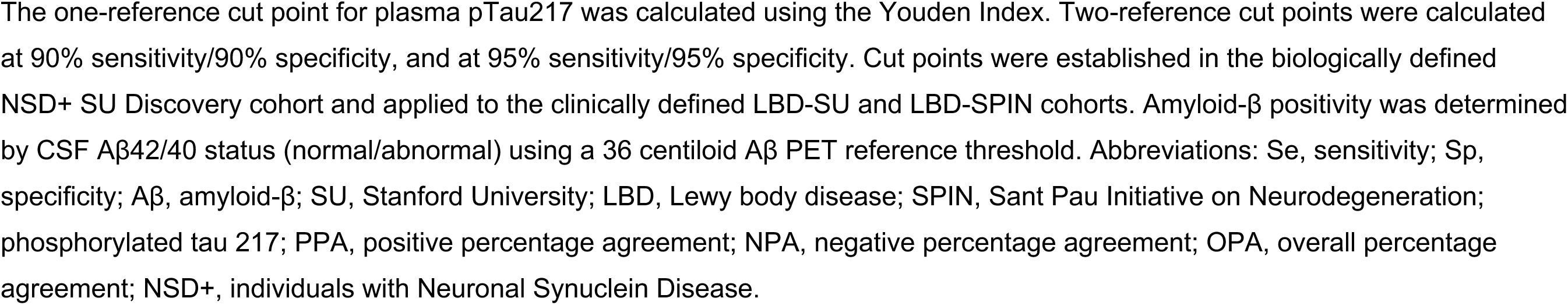
Plasma pTau217 one-and two-reference cut points for amyloid-β positivity in discovery and clinical validation cohorts.

However, the two-reference cut point model significantly improved the positive percentage agreement (PPA) (LBD-SU: 100% vs. 72.7%; LBD-SPIN: 86.4% vs. 68.8%; two-reference cut point vs one-reference cut point, respectively). At 95% sensitivity/specificity, the two-reference cut point model achieved perfect PPA (100%) in both cohorts while maintaining high NPA (LBD-SU: 93.8%, LBD-SPIN: 90.9%).

In the one-reference cut point model, the high NPA but relatively low PPA suggests that plasma pTau217 positive individuals would require additional testing (CSF or PET) to confirm amyloid-β positivity (Figure 3, panel B). With the two-reference cut point at 90% sensitivity/specificity, both intermediate and positive individuals would require additional testing. However, in the 95% sensitivity/specificity model, only intermediate individuals would require additional testing (Figure 3, panel B).

In an exploratory analysis using a 24 CL threshold for Aβ PET positivity, we observed both differences and similarities. The two-reference cut point models significantly improved PPA, reaching 100% in the LBD-SU cohort, while in LBD-SPIN, PPA remained moderate (83.3% in the 90% sensitivity/specificity and 81.3% in the 95% sensitivity/specificity model). NPAs remained consistently high across cohorts for both one-and two-reference cut point models (87.5% to 100%; Supplementary Table 10).

To assess potential impact of using plasma pTau217 on clinical trial recruitment, we calculated the number needed to screen for 100 Aβ+ and or 100 Aβ– NSD participants under each model. The 95% sensitivity/specificity two-cut point model, while offering stringent accuracy, required screening the largest number of patients to find 100 Aβ+ (see Figure 3, panels B and C).

## Discussion

Our findings showed that plasma pTau217 accurately detects amyloid-β in individuals with NSD. To our knowledge, this is the first study analyzing plasma biomarkers of amyloid-β, neuroinflammation, and neurodegeneration in NSD. We identified plasma pTau217 as a reliable biomarker for amyloid-β and applied, for the first time in this population, a two-reference cut point approach similar to that proposed in AD(12,15,52). Since mixed pathologic αSyn and amyloid-β accumulation cannot be diagnosed clinically, accessible biomarkers are critically needed. Plasma pTau217 provides a cost-effective, minimally-invasive alternative to Aβ PET or CSF. This is particularly important as concomitant amyloid-β is common in NSD and is associated with faster functional and cognitive decline(2,3,8,9). Thus, detecting amyloid-β in NSD could guide clinical decisions and inform clinical trial design.

In clinical trials targeting αSyn-related cognitive impairment or dementia, excluding participants with amyloid-β may be necessary. Plasma pTau217 could serve as a prescreening tool reducing the need for additional tests. Conversely, the high frequency of amyloid-β in NSD suggests that anti-amyloid therapies may be appropriate for some individuals. A recent NIH RFA calls for phase 2 clinical trials testing the efficacy of anti-amyloid monoclonal antibodies in DLB with concomitant amyloid-β, expanding the application of targeting Aβ in neurodegenerative diseases beyond AD. Validated tools like plasma pTau217 could efficiently identify eligible participants for such trials(53,54).

In this context, both one-and two-reference cut points at 90% sensitivity/specificity effectively identified individuals without amyloid-β, achieving high NPAs. The two-reference cut point method also demonstrated high PPA, allowing more individuals to avoid invasive or expensive testing compared to the one cut point approach. Additionally, the two-reference cut points were very close: 0.66 pg/mL at 90% sensitivity and 0.69 pg/mL at 90% specificity, reflecting a steep transition between amyloid-β negative and positive participants. This narrow range reduces the effect of small measurement variations and suggests that either threshold could be used depending on whether sensitivity or specificity is prioritized.

Plasma pTau217 may also help stratify trial participants, potentially improving the detection of differential treatment responses. For example, a phase 2A trial of neflamapimod in DLB found that participants with abnormal plasma pTau181 levels did not respond to treatment, leading to the exclusion of biomarker-positive participants in the subsequent phase 2B trial(55,56). These findings suggest that amyloid-β may influence therapeutic responses in NSD and highlight the importance of accounting for mixed pathologies in drug development for neurodegenerative diseases.

In clinical practice, the goal may be ruling-in or ruling-out amyloid-β positivity to guide prognosis and management. More stringent cut points at 95% sensitivity/specificity may be appropriate in this setting, helping clinicians provide better guidance to patients and families regarding prognosis and monitoring.

We also showed that different Aβ PET positivity thresholds influence cut point calculations in NSD. Previous studies suggest that CSF AD biomarker levels in LBD are lower than in AD(31,32), indicating the need for disease-specific cut-points(57). Unlike AD, where amyloid-β burden is consistently high, people with NSD can show a broader spectrum of amyloid-β accumulation, emphasizing a need for tailored biomarker thresholds(58).

Several studies have analyzed plasma pTau217, pTau181, Aβ42/40, GFAP and NfL in clinically diagnosed LBD patients (9,16–20,24,27). Using a biomarker-based definition, our findings demonstrated that pTau217, pTau181, and Aβ42/40 differed between Aβ+ and Aβ− individuals, regardless of NSD status or symptom presence. This is important, as studies have suggested different CSF Aβ and tau biomarker performance in those with and without LBD(53).

We also found elevated plasma GFAP levels in Aβ+ individuals regardless of NSD status, while NfL levels were highest in NSD participants with amyloid-β. Further, both GFAP and NfL levels were associated with the presence of NSD and amyloid-β, suggesting that these markers reflect broader neuroinflammatory or neurodegenerative processes rather than being specific to amyloid-β or αSyn pathology(59).

A key strength of our study is the use of a biologically defined discovery cohort of NSD and Aβ status to assess biomarker performance, combined with external validation of plasma pTau217 in two clinical LBD cohorts. By establishing cut points in a cohort defined using the most validated biomarker for underlying αSyn pathology, we provide thresholds that might be applicable in clinical practice and trials. This design bridges the gap between biomarker-based research frameworks and clinical settings where αSyn status is often unknown, supporting the broader use of our findings. Our study also has limitations. The clinical validation cohorts were relatively small. Additionally, all cohorts consisted of primarily non-Hispanic white individuals, which may limit the generalizability of our findings. Finally, future studies should also evaluate the impact of comorbidities on plasma biomarker levels in NSD.

In summary, our study supports the use of plasma pTau217 as a screening tool for amyloid-β in NSD and in clinically-defined LBD, highlighting a critical mixed-pathology dimension within the biomarkers landscape. Our findings suggest that plasma pTau217 could be used for screening and stratification in NSD clinical trials. These results are important because they integrate clinical and biomarker data in a multi-etiology framework for the diagnosis of mixed neurodegenerative diseases of aging.

## Data Availability

Anonymized data supporting the conclusions of the current study are available to qualified researchers upon reasonable request and is contingent upon completion of Stanford University's data use agreement approval.

## Acknowledgments

The authors would like to express their gratitude to the research participants. We would like to thank the Stanford ADRC Clinical and Imaging Core, and specifically Dr. Michael Zeineh, Dr. Greg Zaharchuk, and Dr. Guido Davidzon for the amyloid-β PET scan reviews.

