## Supplementary Material for "Plasma phosphorylated tau 217 detects amyloid-β in Neuronal Synuclein Disease"

**Supplementary Methods**

### 1. Stanford University (SU) Research Cohorts: SU Discovery and LBD-SU

#### *1.1. Participants*

Both Stanford University (SU) cohorts [SU Discovery (n=180), LBD-SU (n=29)] were comprised of two independent, observational cohorts: one sample from the Iqbal Farrukh and Asad Jamal Stanford Alzheimer's Disease Research Center (ADRC)[1] and another from the Pacific Udall Center (PUC)[2]. SU research participants were recruited between August 2011 and January 2023. All participants or their legally authorized representatives provided written informed consent for participation according to the Declaration of Helsinki. The Institutional Review Board of Stanford University granted approval of the study protocols.

SU research participants are followed annually and undergo detailed clinical evaluations that include comprehensive neuropsychological assessments, collection of blood and CSF, and PET/MR imaging. Clinical diagnoses (LBD-CI, LBD-CU, AD, and CU) were based on international consensus criteria, as described in the manuscript. The study's comprehensive clinical and neuropsychological examination protocols are described elsewhere[1,3,4].

#### *1.2. Blood Collection and Plasma Analysis*

SU research participants consented to donate blood samples during each annual research visit. Ethylenediaminetetraacetic acid (EDTA) plasma was collected by venipuncture, centrifuged for 10 minutes at 4°C at 2000 x *g*, aliquoted in polypropylene tubes, and stored at -80°C until biomarker measurement.

1.2.1. Plasma pTau181 and Plasma Aβ42/40

Plasma pTau181[4] and Aβ42/40[1] were analyzed at the Stanford ADRC Biomarker Core using the fully-automated Lumipulse *G* 1200 platform (Fujirebio Diagnostics, Malvern, PA, USA). The protocol for this analysis has been published previously[1,4,5] and is summarized below.

Plasma pTau181[4,5] samples were analyzed in a single batch in April 2021. Previously non-thawed plasma samples were thawed on wet ice, centrifuged for 5 minutes at 4°C at 500 x *g*, then loaded onto the fully automated Lumipulse *G* 1200 analyzer. Plasma pTau181 levels were quantified using a modified version of the Lumipulse *G* 1200 CSF pTau181 assay (231654; Fujirebio Diagnostics). To reduce the potential for non-specific binding, samples were pre-treated with a heterophilic blocking reagent (200 µg/ml; Scantibodies Inc., Santee, CA, USA)[6]. Samples were measured in singlicate. The inter-batch coefficient of variance (CV) was calculated by retesting 6 independent plasma aliquots with a different batch of reagents and assay lot numbers 1 year after original collection, demonstrating (3.38% variance between the two pTau181 batches). All plasma samples from the present study fell within the quantifiable range of 0.16 to 10.43 pg/ml.

Plasma Aβ42 and Aβ40[1] samples were analyzed in a single batch in May 2023. Previously non-thawed plasma samples were thawed on wet ice, centrifuged for 5 minutes at 4°C at 1000 x *g*, then loaded onto the fully automated Lumipulse *G* 1200 instrument. Within and between batch coefficient of variances (CVs) were calculated by retesting 6 independent plasma aliquots with a different batch of reagents and assay lot numbers 1 year later, demonstrating high test-retest reliability (3.19% variance between the two Aβ42/40 batches). Plasma Aβ42/40 ranged from 0.0549 to 0.2031, with a CV of 2.93%.

1.2.2. Plasma pTau217, GFAP, and NfL

Blood samples were also analyzed at the Department of Psychiatry and Neurochemistry at the University of Gothenburg to determine plasma pTau217, glial fibrillary acidic protein (GFAP), and neurofilament light (NfL) levels. Samples were analyzed in a single batch in December 2023, using the Single molecule array (Simoa®) HD-X platform (Quanterix, Billerica, MA). The University of Gothenburg's protocol for plasma pTau217, GFAP, and NfL analysis has been previously published[7] and is briefly described below.

Previously non-thawed plasma pTau217[7] samples were thawed at room temperature, vortexed for 30 seconds at 2000 rpm, and centrifuged for 10 minutes at 4000 x *g*. Samples were then loaded onto the Quanterix Simoa® HD-X platform, and plasma pTau217 levels were determined using the ALZpath Simoa® pTau217 V2 assay kit (Quanterix, REF: 104371, LOT: 999024). As previously described, the ALZpath pTau217 assay uses a proprietary capture antibody specific to pTau217, an N-terminal detector antibody, and a peptide calibrator. The batch-specific measurement range was 0.0072 to 30.0 pg/mL for SU cohort. Calibrators were run in duplicates, with replicates masked before curve fitting. Samples were diluted three-fold and run in singlicates, with results adjusted for dilution. Three quality-control levels were tested in duplicates at the beginning and end of each run.

Previously non-thawed plasma GFAP and NfL[7] samples were thawed at room temperature, vortexed for 30 seconds at 2000 rpm, and centrifuged for 10 minutes at 4000 x *g*. Samples were then loaded onto the Quanterix Simoa® HD-X platform, and plasma GFAP and NfL levels were determined using the Simoa® Neuro 2-Plex B reagent Kit (Quanterix, REF: 103520, LOT: 503812). One SU participant's GFAP levels fell below the assay's lower limit of quantification and was excluded from statistical analysis. Calibrators were run in duplicates, with replicates masked before curve fitting. Samples were diluted four-fold and run in singlicates, with results adjusted for dilution. Two quality-control levels were tested in duplicates at the beginning and end of each run.

#### *1.3. CSF Collection and Analysis*

Complete protocol for CSF collection and biomarker analysis (CSF αSyn, Aβ42, Aβ40) has been published previously[4,8]. CSF was obtained through lumbar puncture performed at the L4-L5 or L5-S1 interspace using a 20-22 G spinal needle. CSF was collected in externally threaded Thermo Scientific™ Nalgene™ General Long-Term Storage Cryogenic Tubes and were stored in aliquots at -80°C until analysis, with a maximum of two freeze-thaw cycles. All 180 participants in the SU discovery cohort had CSF data available for analysis.

CSF αSyn status was determined using Amprion's αSyn seed amplification assay (SYNTap® Biomarker Test), validated for clinical use under CAP/CLIA for the identification of misfolded αSyn aggregates in CSF. The clinical version of the assay[9] was performed according to standard operational procedures, in agreement in CLIA regulations. Details of the SYNTap® Biomarker test, as well as the protocol used for this study, have been published previously[8]. Briefly, the SYNTap® Biomarker Test amplifies trace amounts of misfolded αSyn aggregates present in CSF to levels that can be measured by thioflavin-T (ThT) fluorescence. CSF samples are combined with buffer containing ThT and recombinant αSyn, then incubated with intermittent shaking cycles until the fluorescence becomes measurable. Each sample was run in triplicate using 40 µL CSF per well in 96-well plates, with reaction mixtures containing 0.3 mg/mL recombinant αSyn, 100 mM PIPES buffer (pH 6.50), 500 mM NaCl, 10 mM ThT, and BSA-blocked borosilicate glass beads in a final volume of 200 µL. Sealed plates underwent baseline fluorescence measurement (440 nm excitation/490 nm emission) before incubation at 37°C with orbital shaking cycles (800 rpm for 1 min followed by 29 min rest) for 7-10 days, with daily fluorescence readings.

CSF samples were classified as αSyn “detected” (NSD+) or αSyn “not detected” (NSD-) based on a preestablished threshold for the median maximum fluorescence of the triplicate wells, within 150 hours. If all three replicates from a sample were positive, CSF samples were deemed “αSyn detected”. If two out of three replicates were positive, samples were deemed inconclusive. Samples with zero or one positive replicate were deemed “αSyn not detected”.

CSF Aβ42 and Aβ40 biomarkers were quantified by the Stanford ADRC Biomarker Core using the Lumipulse *G* 1200 instrument, as previously described[8]. As detailed in the next section, we used Aβ PET as our reference standard to calculate a CSF Aβ42/40 cut point for amyloid-β.

#### *1.4. Aβ PET*

Aβ PET scans were acquired on a PET/MRI scanner (Signa 3 T, GE Healthcare) at the Richard M. Lucas Center for Imaging, Stanford University. Details of image acquisition and processing, are described elsewhere[10]. Briefly, emission data was collected 90-110 minutes after injecting 8.1 milliCuries of $^{18}$F-florabetaben. PET images were reconstructed in 5-minute intervals using standard techniques, then were realigned and summed. FreeSurfer regions of interest (ROIs) from each participant’s structural MRI were used to extract intensity values from the co-registered summed PET data. Standardized uptake value ratios (SUVRs) were calculated for a global cortical ROI using the whole cerebellum as the reference region, then were converted to centiloids (CL) using the Royse et al. equation[11].

To establish a CL cut point for Aβ PET positivity, we analyzed an independent cohort of 72 participants from the Stanford ADRC who were not included in the plasma biomarker analyses. This group had a median (min, max) age of 73 (36-86) years and included 34 females (47.2%) and 38 males (52.8%). Participants were clinically diagnosed with normal cognition (n = 26, 36.1%), Parkinson’s disease without cognitive impairment (*n* = 6, 8.3%), or mild cognitive impairment or dementia due to Alzheimer’s disease (n = 16, 22.2%), Lewy bodies (n = 7, 9.7%), or other etiologies (n = 17, 23.6%: MCI and dementia due to CVD n = 6, FTD n = 1, MCI and dementia due to TBI n = 2, PSP with normal cognition n = 1, MCI due to CBD n = 1, MCI due to PTSD n = 1, MCI due to substance abuse n = 1, MCI due to toxic metabolic encephalopathy n = 1, Cognitive impairment with unknown etiology n = 3).

These participants had a median (min, max) Aβ PET burden of 20.5 [(-13)-188] CL. The distribution of global cortical CL values in this group was modeled using Gaussian finite mixture modeling (GMM) implemented in R (package `mclust`, version 6.1.1). The optimal model identified by the BIC was a two-component GMM with unequal variance. The two Gaussian components corresponded to putative Aβ-negative (mean: 12.8 CL; SD: 10.0 CL) and Aβ-positive (mean: 93.0 CL; SD: 45.2 CL) subpopulations, with estimated proportions of 62% and 38%, respectively. The intersection point between the two fitted distributions was calculated as the value at which their densities were equal, yielding a cut point of 36 CL (Supplementary Figure 2). This threshold was visually confirmed using density plots and applied to the study cohort to classify individuals as Aβ PET positive (CL ≥ 36.) or negative (CL < 36). 47 participants included for GMM analysis were classified Aβ PET-negative (65.3%) and 25 participants were Aβ PET-positive (34.7%).

With Aβ PET as the reference standard, we then used the Youden Index to calculate a CSF Aβ42/40 threshold for amyloid-β (Aβ+) in the SU discovery cohort. The 36CL reference standard yielded a CSF Aβ42/Aβ40 cut point of < 0.09 (AUC = 0.98, 95% CI: 0.95-1). In an exploratory analysis, we also used a threshold of 24 CL from previous studies[12] to favor sensitivity over specificity (eTable1). The 24CL reference standard yielded a CSF Aβ42/40 cut point of < 0.11 (AUC = 0.94, 95% CI: 0.84-1).

The 36CL and 24CL Aβ PET thresholds yielded CSF Aβ42/40 cut points with 89% and 90% sensitivity, respectively, ensuring that approximately 90% of participants with amyloid pathology (as determined by Aβ PET) were correctly classified as CSF Aβ+. Biomarker evidence of amyloid-β was established by abnormal CSF Aβ42/40 ratio, determined by the 36CL Aβ PET threshold, and by the 24CL threshold in our exploratory analyses.

#### *1.5. APOE genotyping*

*APOE* genotype for Stanford University research participants was obtained from National Cell Repository for Alzheimer's Disease (NCRAD) using a Fluidigm fingerprint panel, or determined using PCR restriction fragment length polymorphism analysis within the Stanford ADRC[13]. *APOE* genotype was available for 166 SU Discovery participants and 29 LBD-SU participants.

#### *1.6. MoCA and MMSE Scores*

Global cognition was measured with either the Montreal Cognitive Assessment (MoCA)[14] in 183 participants or the Mini-Mental State Examination (MMSE)[15] in 26 participants (see Table 1, and Supplementary Tables 1 and 2). MMSE scores were converted to MoCA scores according to age, sex, and educational attainment based on Monsell et al.[16] All MoCA and MMSE scores were obtained within a 6-month window of plasma collection.

### 2. Sant Pau Initiative on Neurodegeneration (SPIN)

#### *2.1. Participants*

The SPIN cohort is an observational, multimodal biomarker platform for studying neurodegenerative diseases. Inclusion and exclusion criteria, as well as a comprehensive description of SPIN protocol, are detailed elsewhere[17]. The LBD-SPIN cohort from the present study was recruited between June 2015 and July 2023. All participants or their legally authorized representatives provided written informed consent for participation according to the Declaration of Helsinki. The Institutional Review Board of Hospital Sant Pau granted approval of the study protocol.

SPIN participants consented to undergo comprehensive neurological and neuropsychological evaluation, donate CSF and blood samples, and are followed annually for at least four years. Neuropsychiatric symptoms, functional impact, and the level of global cognitive impairment are also assessed. Mini-Mental State Examination (MMSE)[18,19] scores were included as descriptive indicators of global cognitive status for the present study's LBD-SPIN cohort. All assessments were conducted in Spanish.

The present study included cross-sectional data from 44 SPIN participants clinically diagnosed with mild cognitive impairment (MCI)[20] or dementia due to Lewy bodies (DLB)[21] who also had CSF biomarker confirmation of Aβ status (CSF Aβ42/40).

#### 2.2. Blood Collection and Plasma Analysis

SPIN biofluid samples after neurological and neuropsychological evaluation, during the same study visit. Blood samples were collected in 10 mL EDTA-2K tubes, then centrifuged, aliquoted, and stored at -80°C until analysis. Plasma pTau217 analysis was performed at the Department of Psychiatry and Neurochemistry at the University of Gothenburg using the ALZpath Simoa assay – following a nearly identical protocol as the Stanford cohort. Samples were analyzed in a single batch in March 2023. Instead of three quality-control levels, two quality-control levels were tested in duplicates at the beginning and the end of each run. The batch specific range was 0.0073-30.0 pg/mL.

#### *2.3. CSF Collection and Analysis*

CSF samples were collected in 10 mL polypropylene tubes (Sarstedt, #62.610.018). Within 2 hours, the samples were centrifuged, aliquoted, and stored at -80°C until analysis. CSF Aβ42 and Aβ40 were quantified using the Lumipulse G600II at the Sant Pau Memory Unit laboratory, as previously described[22].

#### *2.4 APOE genotyping*

DNA was extracted from SPIN participants' whole blood samples using the DNeasy® Blood & Tissue kit (Qiagen) at the Sant Pau Memory Unit laboratory. *APOE* genotyping was performed by direct DNA sequencing of exon 4, followed by visual analysis of the electropherograms to identify the two coding polymorphisms (rs429358 and rs7412) that determine the three possible *APOE* isoforms (E2, E3, and E4). 43 out of 44 LBD-SPIN participants had *APOE* genotype data available for analysis.

**Supplementary Figures**

**Supplementary Figure 1.** Selection of research participants

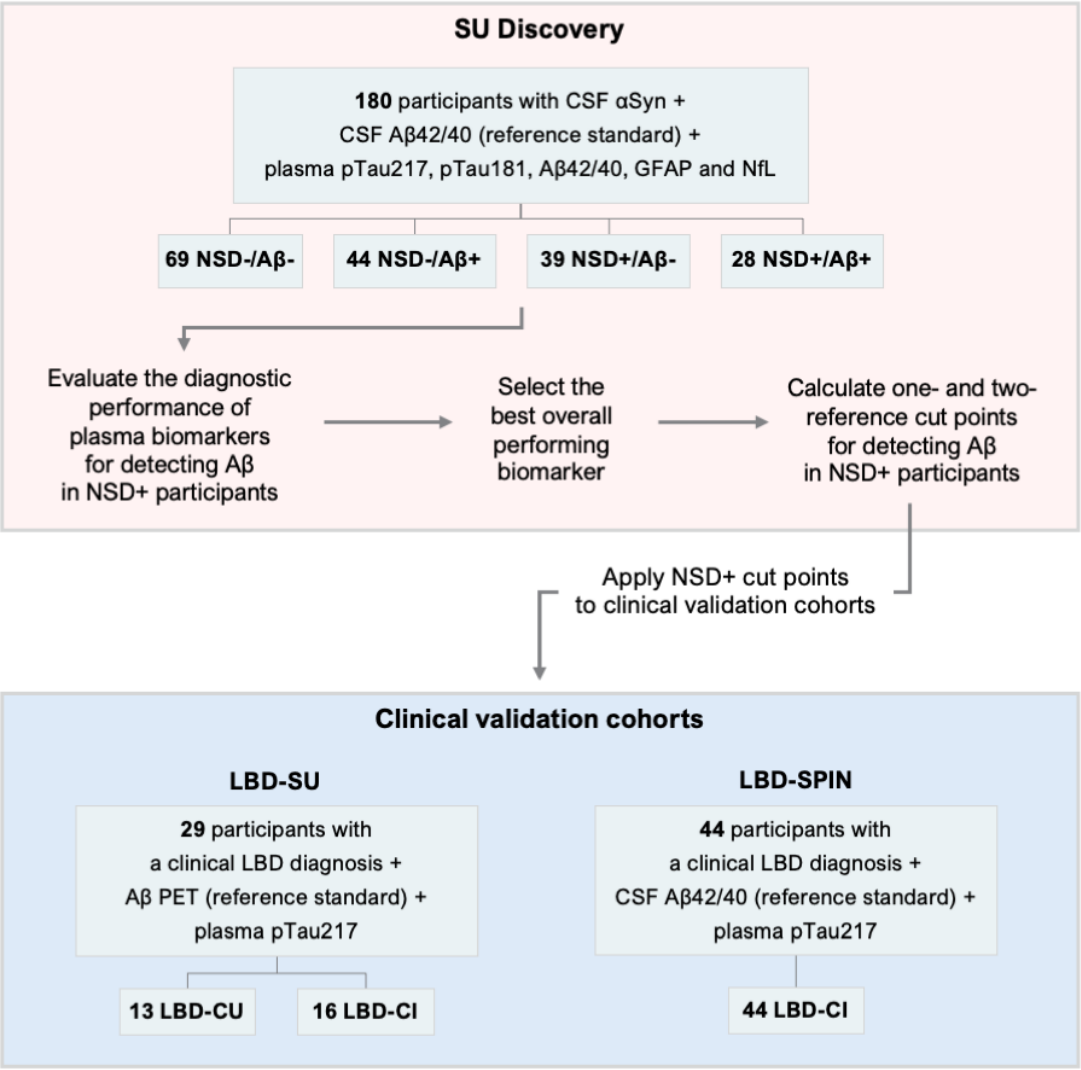

The Stanford University (SU) discovery cohort comprised of 180 biologically-defined research participants of known NSD and Aβ status. In the SU Discovery cohort, we evaluated the diagnostic performance of plasma biomarkers pTau217, pTau181, Aβ42/40, GFAP, and NFL for detecting amyloid-β in NSD+ participants. We then selected the best overall performing biomarker, and calculated one- and two-reference cut points for detecting amyloid-β in NSD+ participants. These cut points were then applied to two independent clinical cohorts LBD-SU and LBD-SPIN, comprised of participants diagnosed with Lewy body disease (LBD) with normal cognition (LBD-CU) or LBD with cognitive impairment (LBD-CI).

**Supplementary Figure 2.** Density plot of Aβ PET centiloid (CL) values in a subset of SU participants excluded from the discovery and validation cohorts

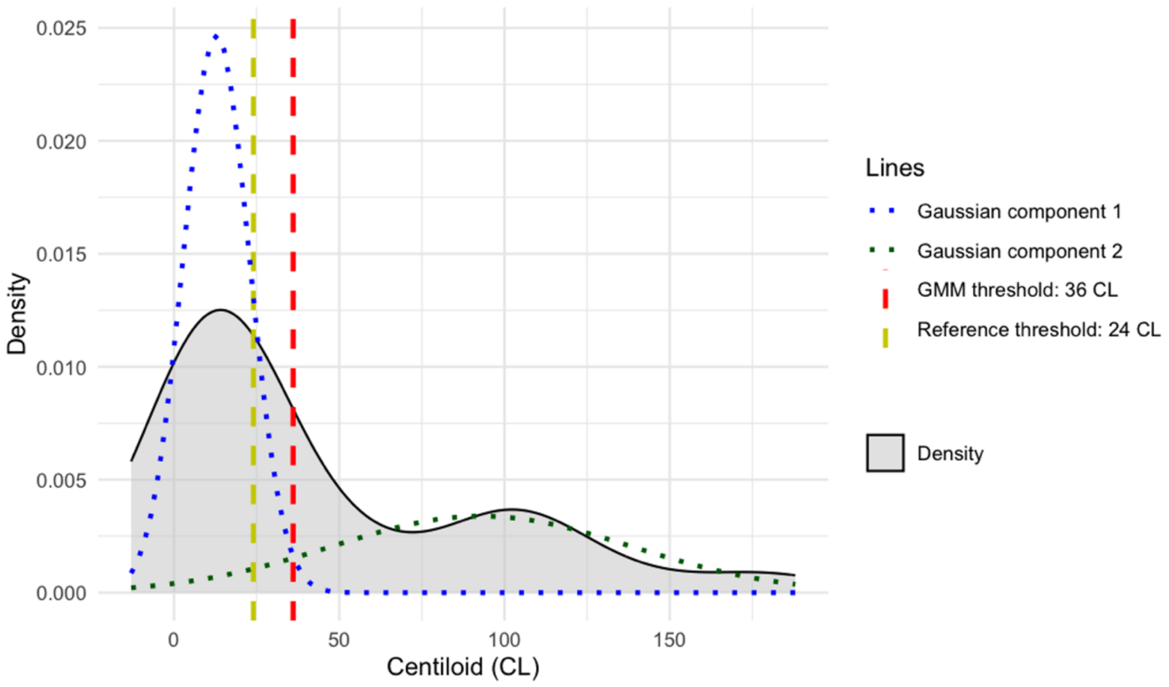

The red dashed line represents the data-driven cut-point at 36 CL, derived using a Gaussian Mixture Model, while the yellow dashed line indicates the reference threshold at 24 CL. The grey shaded area shows the overall density of CL values. The blue and green dotted curves correspond to the two Gaussian components fitted to the data, illustrating the underlying mixture distribution.

**Supplementary Figure 3.** Plasma biomarker levels (pg/mL) across NSD/Aβ groups, stratified by disease status

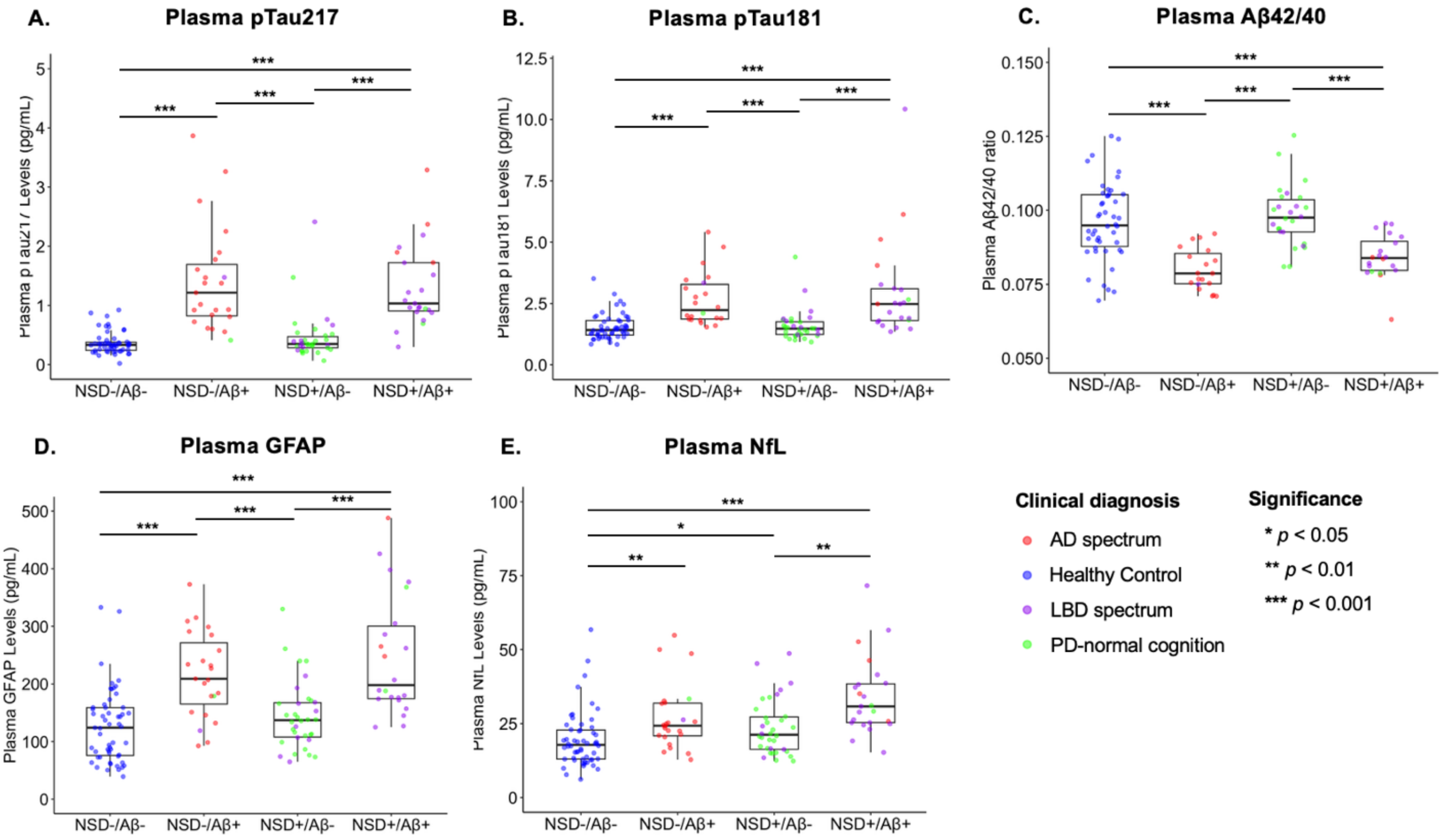

Panel 1A-1E: Bars with asterisks represent significant differences between log transformed plasma biomarker levels. Plasma pTau217, pTau181, Aβ42/40, and GFAP levels were abnormal in Aβ+ groups, regardless of NSD status. Plasma NfL levels were higher in the NSD+/Aβ+ group compared to the NSD-/Aβ- and NSD+/Aβ- groups. Plasma NfL levels were higher in the NSD-/Aβ+ group compared to the NSD-/Aβ- group. Plasma NfL levels were higher in the NSD+/Aβ- group compared to the NSD-/Aβ- group.

**Supplementary Table 1.** Exploratory: Characteristics of the Stanford University discovery cohort using CSF Aβ42/40 <0.11

| | **NSD-/Aβ- (n=60)** | **NSD-/Aβ+ (n=53)** | **NSD+/Aβ- (n=35)** | **NSD+/Aβ+ (n=32)** | **Total Cohort (N=180)** | ***p* value*** |
|---|---|---|---|---|---|---|
| **Age, years** median (min, max) | 66 (51, 87) | 71 (54, 86) | 68 (50, 82) | 71 (57, 83) | 69 (50, 87) | 0.05 |
| **Sex, male** No. (%) | 28 (46.7) | 27 (50.9) | 14 (40.0) | 14 (43.8) | 83 (46.1) | 0.78 |
| **Years of education** median (min, max) | 16 (12, 20) | 16 (5, 20) | 16 (12, 20) | 16 (12, 20) | 16 (5, 20) | 0.98 |
| **APOE ε4 carrier**, No. (%) | 10 (16.7)[b,c,d] | 34 (64.2)[a,c,d] | 10 (28.6)[a,b,d] | 19 (59.4)[a,b,c] | 73 (40.6) | <0.001 |
| **Clinical diagnosis**, No. (%) | | | | | | |
| Cognitively unimpaired | 48 (80.0%)[b,c,d] | 28 (52.8%)[a,c,d] | 4 (11.4%)[a,b,d] | 6 (18.8%)[a,b,c] | 86 (47.8%) | <0.001 |
| AD | 7 (11.7%) | 23 (43.4%) | 0 (0%) | 4 (12.5%) | 34 (18.9%) | |
| LBD Cognitively Impaired | 2 (3.3) | 1 (1.9) | 9 (25.7) | 18 (56.3) | 30 (16.7) | |
| LBD Cognitively Unimpaired | 3 (5.0) | 1 (1.9) | 22 (62.9) | 4 (12.5) | 30 (16.7) | |
| **Clinical status**, No. (%) | | | | | | |
| Asymptomatic | 48 (80.0)[b,c,d] | 28 (52.8)[a,c,d] | 4 (11.4)[a,b] | 6 (18.8)[a,b] | 86 (47.8) | <0.001 |
| Symptomatic | 12 (20.0) | 25 (47.2) | 31 (88.6) | 26 (81.3) | 94 (52.2) | |
| **MoCA score** | 27 (6, 30)[b,d] | 26 (1, 30)[a] | 26 (8, 30)[d] | 22 (3, 30)[a,c] | 26 (1, 30) | 0.001 |
| **CSF Aβ42/40** median (min, max) | 0.13 (0.11, 0.15)[b,d] | 0.07 (0.04, 0.10)[a,c] | 0.12 (0.11, 0.15)[b,d] | 0.07 (0.04, 0.10)[a,c] | 0.11 (0.04, 0.15) | <0.001 |
| **Plasma pTau217, pg/mL** median (min, max) | 0.34 (0.15, 1.88)[b,d] | 0.72 (0.02, 3.87)[a,c] | 0.34 (0.06, 1.47)[b,d] | 0.95 (0.27, 3.29)[a,c] | 0.47 (0.02, 3.87) | <0.001 |
| **Plasma pTau181, pg/mL** median (min, max) | 1.42 (0.83, 3.18)[b,d] | 1.97 (1.09, 5.41)[a,c] | 1.47 (0.93, 4.39)[b,d] | 1.90 (1.18, 10.43)[a,c] | 1.60 (0.83, 10.43) | <0.001 |
| **Plasma GFAP, pg/mL** mean (SD) | 128.76 (64.62)[b,d] | 194.42 (93.14)[a,c] | 141.79 (57.48)[b,d] | 232.81 (121.14)[a,c] | 168.84 (93.09) | <0.001 |
| **Plasma NfL, pg/mL** median (min, max) | 19.18 (6.15, 56.77) | 22.13 (9.87, 54.84) | 20.88 (12.35, 48.72) | 27.91 (15.27, 71.62) | 22.49 (6.15, 71.62) | <0.001 |
| **Plasma Aβ42/40** mean (SD) | 0.10 (0.01)[b,d] | 0.08 (0.01)[a,c] | 0.10 (0.01)[b,d] | 0.08 (0.01)[a,c] | 0.09 (0.01) | <0.001 |

Note. NSD status was determined by CSF αSyn SAA. Clinical status = presence (i.e., symptomatic) or absence (i.e., asymptomatic) of cognitive symptoms; **p* values for descriptive comparisons are uncorrected; [a]$p$<0.05 compared with NSD-/Aβ-; [b]$p$<0.05 compared with NSD-/Aβ+; [c]$p$<0.05 compared with NSD+/Aβ-; [d]$p$<0.05 compared with NSD+/Aβ+. Missing data: APOE ε4 carriership: 14 (7.8%); MoCA: 2 (1.1%); Plasma pTau217: 3 (1.7%); Plasma pTau181: 7 (3.9%); Plasma GFAP and NfL: 1 (0.6%); Plasma Aβ42/40: 32 (17.8%).

Abbreviations: NSD, Neuronal Synuclein Disease; Aβ, amyloid-β; AD, Alzheimer's disease; LBD, Lewy body disease; MoCA, Montreal Cognitive Assessment; CSF, cerebrospinal fluid; pTau217, phosphorylated tau 217; pTau181, phosphorylated tau 181; GFAP, glial fibrillary acidic protein; NfL, neurofilament light chain.

**Supplementary Table 2.** Characteristics of clinical validation cohorts LBD-SU and LBD-SPIN

| | **LBD-SU (N=29)** | **LBD-SPIN (N=44)** |
|---|---|---|
| **Age, years** median (min, max) | 71 (57, 81) | 78 (65, 84) |
| **Sex**, No. (%) | | |
| Male | 6 (20.7) | 20 (45.5) |
| Female | 23 (79.3) | 24 (54.5) |
| **Years of education** median (min, max) | 16 (12, 20) | 8 (0, 20) |
| **APOE ε4 carrier**, No. (%) | | |
| No | 15 (51.7) | 26 (59.1) |
| Yes | 14 (48.3) | 17 (38.6) |
| **Clinical diagnosis**, No. (%) | | |
| LBD Cognitively Unimpaired | 13 (44.8) | - |
| LBD Cognitively Impaired | 16 (55.2) | 44 (100) |
| **Global cognition score** | 25 (15, 30) | 22 (10, 29) |
| **Aβ Status**, No. (%) | | |
| Positive | 20 (69.0) | 21 (47.7) |
| Negative | 9 (31.0) | 23 (52.3) |
| **Plasma pTau217, pg/mL** mean (SD) | 0.54 (0.32) | 0.76 (0.46) |

Note. Global cognition score was determined by MoCA in LBD-SU and MMSE in LBD-SPIN. Aβ Status was determined by Aβ PET in LBD-SU and CSF Aβ42/40 in LBD-SPIN. Missing data: Years of education: 3 (6.8%); APOE ε4 carriership: 1 (2.4%); MoCA: 4 (13.8%); MMSE: 9 (20.5%); Plasma pTau217: 3 (1.7%).

Abbreviations: LBD, Lewy body disease; SU, Stanford University; SPIN, Sant Pau Initiative on Neurodegeneration; Aβ, amyloid-β; MoCA, Montreal Cognitive Assessment; MMSE, Mini Mental State Examination; phosphorylated tau 217.

**Supplementary Table 3.** Median fold-change of plasma biomarkers in NSD+ and NSD- participants

| | NSD+ | | | NSD- | | |
|---|---|---|---|---|---|---|
| | **Median Fold-Change** | **95% CI** | **SD** | **Median Fold-Change** | **95% CI** | **SD** |
| **Plasma pTau181** | 1.36 | 1.12-1.85 | 0.21 | 1.41 | 1.27-1.67 | 0.10 |
| **Plasma pTau217** | 2.79 | 2.30-3.61 | 0.33 | 2.49 | 2.06-3.52 | 0.35 |
| **Plasma Aβ42/40** | 1.17 | 0.80-0.90 | 0.02 | 1.19 | 0.78-0.89 | 0.03 |
| **Plasma GFAP** | 1.50 | 1.25-2.20 | 0.26 | 1.68 | 1.25-2.33 | 0.27 |
| **Plasma NfL** | 1.34 | 1.09-1.67 | 0.15 | 1.25 | 1.06-1.42 | 0.09 |

Note. Median fold-change in plasma biomarker levels was determined between Aβ+ and Aβ- participants, across NSD+ and NSD- groups. Associated 95% confidence intervals [CI] and standard deviation [SD] are also reported.

Abbreviations: NSD+, individuals with Neuronal Synuclein Disease; NSD-, individuals without Neuronal Synuclein Disease; pTau181, phosphorylated tau 181; pTau217, phosphorylated tau 217; Aβ, amyloid-β; GFAP, glial fibrillary acidic protein; NfL, neurofilament light chain.

**Supplementary Table 4a.** Univariable and multivariable linear regression models for plasma biomarkers

| | **Plasma pTau217** | | | | | **Plasma pTau181** | | | | | **Plasma Aβ42/40** | | | |
|---|---|---|---|---|---|---|---|---|---|---|---|---|---|---|
| **Univariable Regression Models** | **b** | **SE** | **t** | ***p*** | **95% CI** | **b** | **SE** | **t** | ***p*** | **95% CI** | **SE** | **t** | ***p*** | **95% CI** |
| Predictor 1 | | | | | | | | | | | | | | |
| Intercept | -0.26 | 0.03 | -10.19 | <0.001 | (-0.31, -0.21) | 0.25 | 0.01 | 18.51 | <0.001 | (0.26, 0.28) | 0.001 | 77.27 | <0.001 | (0.08, 0.09) |
| **Age** | 0.01 | 0.003 | 2.78 | **<0.01** | (0.003, 0.02) | 0.005 | 0.002 | 2.79 | **<0.01** | (0.001, 0.008) | 0.0001 | -1.40 | 0.16 | (-0.0005, 0.0001) |
| Predictor 2 | | | | | | | | | | | | | | |
| Intercept | -0.30 | 0.04 | -8.13 | <0.001 | (-0.37, -0.23) | 0.23 | 0.02 | 11.86 | <0.001 | (0.19, 0.27) | 0.002 | 56.55 | <0.001 | (0.088, 0.09) |
| **Sex Male** | 0.04 | 0.05 | 0.89 | 0.38 | (-0.054, 0.143) | 0.02 | 0.03 | 0.84 | 0.40 | (-0.031, 0.076) | 0.002 | -0.57 | 0.57 | (-0.006, 0.003) |
| Predictor 3 | | | | | | | | | | | | | | |
| Intercept | -0.35 | 0.03 | -10.25 | <0.001 | (-0.42, -0.29) | 0.21 | 0.02 | 11.45 | <0.001 | (0.17, 0.25) | 0.001 | 66.06 | <0.001 | (0.09, 0.10) |
| **APOE ε4 Carrier** | 0.18 | 0.05 | 3.47 | **<0.001** | (0.08, 0.28) | 0.08 | 0.03 | 2.94 | **<0.01** | (0.03, 0.14) | 0.002 | -4.40 | **<0.001** | (-0.01, -0.005) |
| Predictor 4 | | | | | | | | | | | | | | |
| Intercept | -0.46 | 0.02 | -18.94 | <0.001 | (-0.51, -0.41) | 0.17 | 0.01 | 11.65 | <0.001 | (0.14, 0.20) | 0.001 | 77.35 | <0.001 | (0.09, 0.10) |
| **CSF Aβ+** | 0.45 | 0.04 | 11.82 | **<0.001** | (0.38, 0.53) | 0.18 | 0.024 | 7.35 | **<0.001** | (0.13, 0.22) | 0.002 | -77.53 | **<0.001** | (-0.02, -0.01) |
| Predictor 5 | | | | | | | | | | | | | | |
| Intercept | -0.30 | 0.03 | -9.43 | <0.001 | (-0.36, -0.24) | 0.24 | 0.02 | 13.86 | <0.001 | (0.20, 0.27) | 0.001 | 63.10 | <0.001 | (0.087, 0.09) |
| **CSF NSD+** | 0.06 | 0.05 | 1.14 | 0.26 | (-0.04, 0.16) | 0.02 | 0.03 | 0.58 | 0.56 | (-0.04, 0.07) | 0.002 | 0.41 | 0.68 | (-0.004, 0.006) |
| **Multivariable Regression Models** | **b** | **SE** | **t** | ***p*** | **95% CI** | **b** | **SE** | **t** | ***p*** | **95% CI** | **SE** | **t** | ***p*** | **95% CI** |
| Intercept | -0.52 | 0.04 | -12.68 | <0.001 | (-0.60, 0.44) | 0.15 | 0.03 | 6.01 | <0.001 | (0.10, 0.20) | 0.002 | 48.87 | <0.001 | (0.09, 0.10) |
| **Age** | 0.001 | 0.00 | 0.44 | 0.66 | (-0.004, 0.007) | 0.002 | 0.002 | 1.14 | 0.26 | (-0.001, 0.005) | 0.0001 | 0.57 | 0.57 | (-0.0002, 0.0003) |
| **Sex Male** | 0.06 | 0.04 | 1.46 | 0.15 | (-0.02, 0.14) | 0.03 | 0.03 | 1.24 | 0.22 | (-0.02, 0.08) | 0.002 | -1.17 | 0.24 | (-0.006, 0.002) |
| **APOE ε4 Carrier** | 0.02 | 0.04 | 0.55 | 0.58 | (-0.06, 0.11) | 0.02 | 0.03 | 0.86 | 0.39 | (-0.03, 0.08) | 0.002 | -2.41 | **<0.05** | (-0.009, -0.0009) |
| **CSF Aβ+** | 0.45 | 0.04 | 10.34 | **<0.001** | (0.37, 0.54) | 0.17 | 0.03 | 6.15 | **<0.001** | (0.11, 0.22) | 0.002 | -5.97 | **<0.001** | (-0.02, -0.009) |
| **CSF NSD+** | 0.04 | 0.04 | 1.11 | 0.27 | (-0.03, 0.12) | 0.01 | 0.03 | 0.36 | 0.72 | (-0.04, 0.06) | 0.002 | 1.65 | 0.10 | (-0.0007, 0.007) |

Abbreviations: pTau217, phosphorylated tau 217; pTau181, phosphorylated tau 181; Aβ, amyloid-β; CSF, cerebrospinal fluid; CSF Aβ+, individuals with abnormal CSF Aβ42/40 levels; NSD+, individuals with Neuronal Synuclein Disease.

**Supplementary Table 4b.** Univariable and multivariable linear regression models for plasma biomarkers

| | Plasma GFAP | | | | | Plasma NfL | | | | |
|---|---|---|---|---|---|---|---|---|---|---|
| **Univariable Regression Models** | **b** | **SE** | **t** | ***p*** | **95% CI** | **b** | **SE** | **t** | ***p*** | **95% CI** |
| Predictor 1 | | | | | | | | | | |
| Intercept | 2.19 | 0.02 | 130.54 | <0.001 | (2.15-2.22) | 1.37 | 0.01 | 108.48 | <0.001 | (1.34, 1.39) |
| **Age** | 0.01 | 0.002 | 4.17 | **<0.001** | (0.005-0.01) | 0.01 | 0.002 | 6.04 | **<0.001** | (0.007, 0.01) |
| Predictor 2 | | | | | | | | | | |
| Intercept | 2.20 | 0.03 | 88.36 | <0.001 | (2.15, 2.25) | 1.35 | 0.02 | 68.32 | <0.001 | (1.31, 1.39) |
| **Sex Male** | -0.05 | 0.03 | -1.54 | 0.13 | (-0.12, 0.01) | -0.005 | 0.03 | -0.18 | 0.86 | (-0.06, 0.05) |
| Predictor 3 | | | | | | | | | | |
| Intercept | 2.14 | 0.02 | 92.80 | <0.001 | (2.09, 2.18) | 1.33 | 0.02 | 72.94 | <0.001 | (1.30, 1.37) |
| **APOE ε4 Carrier** | 0.08 | 0.04 | 2.25 | **<0.05** | (0.009, 0.15) | 0.03 | 0.03 | 1.11 | 0.27 | (-0.025, 0.086) |
| Predictor 4 | | | | | | | | | | |
| Intercept | 2.08 | 0.02 | 108.7 | <0.001 | (2.04, 2.12) | 1.30 | 0.02 | 79.46 | <0.001 | (1.27, 1.34) |
| **CSF Aβ+** | 0.22 | 0.03 | 7.4 | **<0.001** | (0.16, 0.28) | 0.11 | 0.03 | 4.24 | **<0.001** | (0.06, 0.16) |
| Predictor 5 | | | | | | | | | | |
| Intercept | 2.14 | 0.02 | 100.77 | <0.001 | (2.10, 2.18) | 1.32 | 0.02 | 79.95 | <0.001 | (1.28-1.35) |
| **CSF NSD+** | 0.07 | 0.03 | 2.06 | **<0.05** | (0.003, 0.14) | 0.08 | 0.03 | 3.13 | **<0.01** | (0.03, 0.14) |
| **Multivariable Regression Models** | **b** | **SE** | **t** | ***p*** | **95% CI** | **b** | **SE** | **t** | ***p*** | **95% CI** |
| Intercept | 2.12 | 0.03 | 70.29 | <0.001 | (2.06, 2.18) | 1.31 | 0.02 | 54.74 | <0.001 | (1.26, 1.36) |
| **Age** | 0.01 | 0.002 | 3.07 | **<0.01** | (0.002, 0.01) | 0.01 | 0.002 | 5.21 | **<0.001** | (0.005, 0.01) |
| **Sex Male** | -0.08 | 0.03 | -2.51 | **<0.05** | (-0.14, -0.02) | -0.04 | 0.02 | -1.54 | 0.12 | (-0.08, 0.01) |
| **APOE ε4 Carrier** | 0.00 | 0.03 | 0.13 | 0.90 | (-0.06, 0.07) | -0.01 | 0.03 | -0.36 | 0.72 | (-0.06, 0.04) |
| **CSF Aβ+** | 0.20 | 0.03 | 6.04 | **<0.001** | (0.13, 0.26) | 0.09 | 0.03 | 3.57 | **<0.001** | (0.04, 0.14) |
| **CSF NSD+** | 0.06 | 0.03 | 2.03 | **<0.05** | (0.002, 0.12) | 0.09 | 0.02 | 3.85 | **<0.001** | (0.05, 0.14) |

Abbreviations: GFAP, glial fibrillary acidic protein; NfL, neurofilament light chain; CSF, cerebrospinal fluid; Aβ, amyloid-β; CSF Aβ+, individuals with abnormal CSF Aβ42/40 levels; NSD+, individuals with Neuronal Synuclein Disease.

**Supplementary Table 5.** ROC models of plasma biomarkers for detecting amyloid-β positivity defined by CSF Aβ42/40 <0.09 in NSD+ participants

| Biomarker Model | AUC | 95% CI | Sensitivity | Specificity | *p* value* |
|---|---|---|---|---|---|
| pTau217 | 0.92 | 0.85-1 | 0.96 | 0.85 | - |
| pTau181 | 0.79 | 0.69-0.90 | 0.78 | 0.67 | 0.92 |
| Aβ42/40 | 0.90 | 0.81-0.98 | 0.76 | 0.93 | 0.63 |
| GFAP | 0.82 | 0.72-0.92 | 0.75 | 0.82 | 0.07 |
| NfL | 0.72 | 0.59-0.84 | 0.79 | 0.67 | 0.003 |
| pTau181 + pTau217 | 0.92 | 0.84-1 | 0.93 | 0.92 | 0.95 |
| pTau181 + Aβ42/40 | 0.93 | 0.86-0.99 | 0.88 | 0.83 | 0.96 |
| pTau181 + GFAP | 0.84 | 0.74-0.93 | 0.78 | 0.79 | 0.17 |
| pTau217 + Aβ42/40 | 0.94 | 0.88-1 | 0.92 | 0.87 | 0.75 |
| pTau217 + GFAP | 0.92 | 0.86-0.99 | 0.86 | 0.92 | 0.88 |
| Aβ42/40 + GFAP | 0.94 | 0.89-1 | 0.84 | 0.93 | 0.66 |
| pTau181 + GFAP + NfL | 0.83 | 0.74-0.93 | 0.81 | 0.77 | 0.16 |
| pTau217 + GFAP + NfL | 0.92 | 0.85-0.99 | 0.82 | 0.95 | 0.89 |
| Aβ42/40 + GFAP + NfL | 0.95 | 0.90-1 | 0.88 | 0.93 | 0.54 |
| pTau181 + pTau217 + GFAP + NfL | 0.93 | 0.86-1 | 0.85 | 0.95 | 0.93 |
| pTau181 + Aβ42/40 + GFAP + NfL | 0.96 | 0.92-1 | 1.00 | 0.80 | 0.39 |
| pTau217 + Aβ42/40 + GFAP + NfL | 0.96 | 0.92-1 | 0.92 | 0.90 | 0.37 |
| pTau181 + pTau217 + Aβ42/40 + GFAP + NfL | 0.96 | 0.92-1 | 0.80 | 0.97 | 0.36 |

Note. *DeLong test p-value comparing the ROC AUC of the biomarker model to the ROC AUC of plasma pTau217.

Abbreviations: ROC AUC, receiver operating characteristic area under the curve; SU, Stanford University; pTau217, phosphorylated tau 217; pTau181, phosphorylated tau 181; Aβ, amyloid-β; GFAP, glial fibrillary acidic protein; NfL, neurofilament light chain; CSF, cerebrospinal fluid; NSD+, individuals with Neuronal Synuclein Disease.

**Supplementary Table 6.** ROC models of plasma biomarkers for detecting amyloid-β positivity defined by CSF Aβ42/40 <0.09 in NSD- participants

| Biomarker Model | AUC | 95% CI | Sensitivity | Specificity | *p* value* |
|---|---|---|---|---|---|
| pTau217 | 0.91 | 0.86-0.97 | 0.93 | 0.75 | - |
| pTau181 | 0.80 | 0.72-0.88 | 0.75 | 0.78 | 0.03 |
| Aβ42/40 | 0.83 | 0.75-0.92 | 0.78 | 0.82 | 0.14 |
| GFAP | 0.77 | 0.69-0.86 | 0.72 | 0.71 | 0.01 |
| NfL | 0.66 | 0.56-0.76 | 0.77 | 0.58 | <0.0001 |
| pTau181 + pTau217 | 0.91 | 0.85-0.97 | 0.82 | 0.85 | 0.96 |
| pTau181 + Aβ42/40 | 0.86 | 0.78-0.95 | 0.89 | 0.77 | 0.33 |
| pTau181 + GFAP | 0.84 | 0.76-0.91 | 0.90 | 0.65 | 0.12 |
| pTau217 + Aβ42/40 | 0.92 | 0.86-0.99 | 0.95 | 0.83 | 0.79 |
| pTau217 + GFAP | 0.91 | 0.85-0.97 | 0.83 | 0.85 | 0.97 |
| Aβ42/40 + GFAP | 0.88 | 0.80-0.96 | 0.83 | 0.87 | 0.47 |
| pTau181 + GFAP + NfL | 0.84 | 0.77-0.92 | 0.72 | 0.85 | 0.15 |
| pTau217 + GFAP + NfL | 0.92 | 0.87-0.97 | 0.90 | 0.82 | 0.87 |
| Aβ42/40 + GFAP + NfL | 0.88 | 0.81-0.96 | 0.83 | 0.87 | 0.56 |
| pTau181 + pTau217 + GFAP + NfL | 0.92 | 0.86-0.97 | 0.92 | 0.83 | 0.84 |
| pTau181 + Aβ42/40 + GFAP + NfL | 0.90 | 0.82-0.97 | 0.91 | 0.82 | 0.73 |
| pTau217 + Aβ42/40 + GFAP + NfL | 0.92 | 0.86-0.99 | 0.86 | 0.92 | 0.78 |
| pTau181 + pTau217 + Aβ42/40 + GFAP + NfL | 0.93 | 0.87-1.00 | 0.88 | 0.92 | 0.59 |

Note. *DeLong test *p*-value comparing the ROC AUC of the biomarker model to the ROC AUC of plasma pTau217 alone.

Abbreviations: ROC AUC, receiver operating characteristic area under the curve; SU, Stanford University; pTau217, phosphorylated tau 217; pTau181, phosphorylated tau 181; Aβ, amyloid-β; GFAP, glial fibrillary acidic protein; NfL, neurofilament light chain; CSF, cerebrospinal fluid; NSD-, individuals without Neuronal Synuclein Disease.

**Supplementary Table 7.** ROC models of plasma biomarkers for detecting amyloid-β positivity defined by CSF Aβ42/40 <0.11 in NSD+ participants

| Biomarker Model | AUC | 95% CI | Sensitivity | Specificity | *p* value* |
|---|---|---|---|---|---|
| pTau217 | 0.92 | 0.84-0.99 | 0.94 | 0.83 | - |
| pTau181 | 0.77 | 0.66-0.88 | 0.74 | 0.69 | 0.04 |
| Aβ42/40 | 0.85 | 0.75-0.95 | 0.68 | 0.93 | 0.29 |
| GFAP | 0.77 | 0.66-0.89 | 0.69 | 0.83 | 0.01 |
| NfL | 0.73 | 0.61-0.85 | 0.78 | 0.71 | 0.005 |
| pTau181 + pTau217 | 0.91 | 0.84-0.99 | 0.84 | 0.94 | 0.95 |
| pTau181 + Aβ42/40 | 0.88 | 0.79-0.97 | 0.79 | 0.85 | 0.53 |
| pTau181 + GFAP | 0.79 | 0.68-0.90 | 0.71 | 0.80 | 0.06 |
| pTau217 + Aβ42/40 | 0.95 | 0.89-1 | 0.89 | 0.96 | 0.54 |
| pTau217 + GFAP | 0.92 | 0.84-0.99 | 0.88 | 0.89 | 0.93 |
| Aβ42/40 + GFAP | 0.92 | 0.85-0.99 | 0.79 | 0.96 | 0.94 |
| pTau181 + GFAP + NfL | 0.78 | 0.67-0.89 | 0.87 | 0.57 | 0.05 |
| pTau217 + GFAP + NfL | 0.92 | 0.84-0.99 | 0.84 | 0.94 | 0.93 |
| Aβ42/40 + GFAP + NfL | 0.93 | 0.86-0.99 | 0.75 | 1.00 | 0.83 |
| pTau181 + pTau217 + GFAP + NfL | 0.91 | 0.83-0.99 | 0.84 | 0.94 | 0.94 |
| pTau181 + Aβ42/40 + GFAP + NfL | 0.94 | 0.89-1 | 0.86 | 0.93 | 0.60 |
| pTau217 + Aβ42/40 + GFAP + NfL | 0.95 | 0.90-1 | 0.89 | 0.96 | 0.44 |
| pTau181 + pTau217 + Aβ42/40 + GFAP + NfL | 0.96 | 0.92-1 | 0.89 | 0.96 | 0.31 |

Note. *DeLong test *p*-value comparing the ROC AUC of the biomarker model to the ROC AUC of plasma pTau217 alone.

Abbreviations: ROC AUC, receiver operating characteristic area under the curve; SU, Stanford University; pTau217, phosphorylated tau 217; pTau181, phosphorylated tau 181; Aβ, amyloid-β; GFAP, glial fibrillary acidic protein; NfL, neurofilament light chain; CSF, cerebrospinal fluid; NSD+, individuals with Neuronal Synuclein Disease.

**Supplementary Table 8.** ROC models of plasma biomarkers for detecting amyloid-β positivity defined by CSF Aβ42/40 <0.11 in NSD- participants

| Biomarker Model | AUC | 95% CI | Sensitivity | Specificity | *p* value* |
|---|---|---|---|---|---|
| pTau217 | 0.83 | 0.75-0.91 | 0.83 | 0.74 | - |
| pTau181 | 0.77 | 0.68-0.86 | 0.69 | 0.81 | 0.33 |
| Aβ42/40 | 0.81 | 0.72-0.90 | 0.81 | 0.76 | 0.78 |
| GFAP | 0.72 | 0.64-0.83 | 0.69 | 0.70 | 0.13 |
| NfL | 0.57 | 0.47-0.68 | 0.65 | 0.53 | 0.0002 |
| pTau181 + pTau217 | 0.82 | 0.74-0.91 | 0.73 | 0.86 | 0.90 |
| pTau181 + Aβ42/40 | 0.85 | 0.76-0.93 | 0.88 | 0.76 | 0.77 |
| pTau181 + GFAP | 0.80 | 0.72-0.89 | 0.83 | 0.69 | 0.67 |
| pTau217 + Aβ42/40 | 0.88 | 0.81-0.96 | 0.86 | 0.85 | 0.35 |
| pTau217 + GFAP | 0.83 | 0.75-0.91 | 0.74 | 0.86 | 0.97 |
| Aβ42/40 + GFAP | 0.86 | 0.78-0.94 | 0.76 | 0.90 | 0.63 |
| pTau181 + GFAP + NfL | 0.83 | 0.75-0.91 | 0.79 | 0.79 | 0.97 |
| pTau217 + GFAP + NfL | 0.86 | 0.78-0.93 | 0.82 | 0.88 | 0.61 |
| Aβ42/40 + GFAP + NfL | 0.86 | 0.78-0.94 | 0.76 | 0.90 | 0.63 |
| pTau181 + pTau217 + GFAP + NfL | 0.87 | 0.79-0.94 | 0.80 | 0.88 | 0.50 |
| pTau181 + Aβ42/40 + GFAP + NfL | 0.89 | 0.81-0.96 | 0.90 | 0.82 | 0.29 |
| pTau217 + Aβ42/40 + GFAP + NfL | 0.91 | 0.84-0.98 | 0.85 | 0.92 | 0.15 |
| pTau181 + pTau217 + Aβ42/40 + GFAP + NfL | 0.91 | 0.84-0.98 | 0.90 | 0.92 | 0.13 |

Note. *DeLong test *p*-value comparing the ROC AUC of the biomarker model to the ROC AUC of plasma pTau217 alone.

Abbreviations: ROC AUC, receiver operating characteristic area under the curve; SU, Stanford University; pTau217, phosphorylated tau 217; pTau181, phosphorylated tau 181; Aβ, amyloid-β; GFAP, glial fibrillary acidic protein; NfL, neurofilament light chain; CSF, cerebrospinal fluid; NSD-, individuals without Neuronal Synuclein Disease.

**Supplementary Table 9.** Plasma pTau217 reference cut points for amyloid-β positivity in NSD- participants

| | One cut point (Youden Index) | | Two cut points 90% Se/Sp | Two cut points 95% Se/Sp |
|---|---|---|---|---|
| | pTau217 positive >0.41 pg/mL | | pTau217 positive >0.65 pg/mL pTau217 negative <0.42 pg/mL) | pTau217 positive >0.81 pg/mL pTau217 negative <0.37 pg/mL) |
| **Plasma Aβ positive**, No. (%) | 57 (51.8) | **Plasma Aβ positive**, No. (%) | 38 (34.5) | 26 (23.6) |
| | | **Plasma Aβ intermediate**, No. (%) | 18 (16.4) | 35 (31.8) |
| **Plasma Aβ negative**, No. (%) | 53 (48.2) | **Plasma Aβ negative**, No. (%) | 54 (49.1) | 49 (44.5) |
| **Sensitivity**, % | 93.0 | **Sensitivity, lower cut point**, % | 88.6 | 88.0 |
| **Specificity**, % | 74.6 | **Specificity, upper cut point**, % | 87.7 | 92.0 |
| **PPA**, % | 70.2 | **PPA, upper cut point**, % | 81.6 | 84.6 |
| **NPA**, % | 94.3 | **NPA, lower cut point**, % | 92.6 | 93.9 |
| **OPA**, % | 81.8 | **OPA for plasma pTau217 positive and negative**, % | 88.0 | 90.7 |

Note. The one-reference cut point for plasma pTau217 was calculated using the Youden Index. Two-reference cut points were calculated at 90% sensitivity/90% specificity, and at 95% sensitivity/95% specificity. Cut points were established in the biologically defined NSD- SU discovery cohort. Amyloid-β positivity was determined with CSF Aβ42/40 status (normal/abnormal) using a 36 CL Aβ PET reference threshold.

Abbreviations: Se, sensitivity; Sp, specificity; Aβ, amyloid-β; SU, Stanford University; phosphorylated tau 217; PPA, Positive percentage agreement; NPA, Negative percentage agreement; OPA, overall percentage agreement; PET, positron emission tomography; CSF, cerebrospinal fluid; NSD-, individuals without Neuronal Synuclein Disease.

**Supplementary Table 10.** Plasma pTau217 reference cut points for amyloid-β positivity defined by CSF Aβ42/40 <0.11 in NSD+ participants

| | **One cut point (Youden Index)** | | **Two cut points 90% Se/Sp** | | | **Two cut points 95% Se/Sp** | | |
|---|---|---|---|---|---|---|---|---|
| | pTau217 positive >0.48 pg/mL | | pTau217 positive >0.67 pg/mL pTau217 negative <0.52 pg/mL) | | | pTau217 positive >0.80 pg/mL pTau217 negative <0.33 pg/mL) | | |
| **Cohort** | **SU discovery (NSD+)** | **Cohort** | **SU discovery (NSD+)** | **LBD-SU** | **LBD-SPIN** | **SU discovery (NSD+)** | **LBD-SU** | **LBD-SPIN** |
| **Plasma Aβ positive**, No. (%) | 36 (53.7) | **Plasma Aβ positive**, No. (%) | 29 (43.3) | 6 (20.7) | 11 (25.0) | 22 (32.8) | 6 (20.7) | 16 (36.4) |
| | | **Plasma Aβ intermediate**, No. (%) | 5 (7.5) | 7 (24.1) | 8 (18.2) | 27 (40.3) | 15 (51.7) | 21 (47.7) |
| **Plasma Aβ negative**, No. (%) | 31 (46.3) | **Plasma Aβ negative**, No. (%) | 33 (49.3) | 16 (55.2) | 24 (54.5) | 18 (26.9) | 8 (27.6) | 7 (15.9) |
| **Sensitivity**, % | 93.8 | **Sensitivity, lower cut point**, % | 89.7 | 75.0 | 95.2 | 91.3 | 85.7 | 100.0 |
| **Specificity**, % | 82.9 | **Specificity, upper cut point**, % | 90.9 | 100.0 | 71.4 | 94.1 | 100.0 | 70.0 |
| **PPA**, % | 83.3 | **PPA, upper cut point**, % | 89.7 | 100.0 | 83.3 | 95.5 | 100.0 | 81.3 |
| **NPA**, % | 93.6 | **NPA, lower cut point**, % | 90.9 | 87.5 | 90.9 | 88.9 | 87.5 | 100.0 |
| **OPA**, % | 88.1 | **OPA for plasma pTau217 positive and negative**, % | 90.3 | 90.9 | 85.7 | 92.5 | 92.9 | 86.96 |

Note. The one-reference cut point for plasma pTau217 was calculated using the Youden Index. Two-reference cut points were calculated at 90% sensitivity/90% specificity, and at 95% sensitivity/95% specificity. Cut points were established in the biologically defined NSD+ SU discovery cohort and applied to the clinically defined LBD-SU and LBD-SPIN cohorts. Amyloid-β positivity was determined with CSF Aβ42/40 status (normal/abnormal) using a 24 CL Aβ PET reference threshold.

Abbreviations: Se, sensitivity; Sp, specificity; Aβ, amyloid-β; SU, Stanford University; LBD, Lewy body disease; SPIN, Sant Pau Initiative on Neurodegeneration; phosphorylated tau 217; PPA, Positive percentage agreement; NPA, Negative percentage agreement; OPA, overall percentage agreement; PET, positron emission tomography; CSF, cerebrospinal fluid; NSD+, individuals with Neuronal Synuclein Disease.
